# Analysis of tweets discussing the risk of Mpox among children and young people in school (May-Oct 2022): Public health experts on Twitter consistently exaggerated risks and infrequently reported accurate information

**DOI:** 10.1101/2023.05.11.23289839

**Authors:** Benjamin Knudsen, Tracy Beth Høeg, Vinay Prasad

## Abstract

**Importance:** Twitter is used by health professionals to relay information. We sought to investigate the use of tweets to describe Mpox risks to children and young people in school during summer/ fall of 2022.

**Objective:** To determine the number of tweets discussing the risk of Mpox to children and young people in school and 1) determine accuracy, 2) for inaccurate tweets, determine if risk was minimized or exaggerated and 3) describe the characteristics of the accounts and tweets which contained accurate vs. inaccurate information.

**Design:** Retrospective observational study.

**Setting:** Twitter advanced search in January 2023 of tweets spanning May 18th, 2022, to September 19th, 2022.

**Participants:** Accounts labeled as: MD, DO, nurse, pharmacist, physical therapist, other health care provider, PhD, MPH, other Ed. degree, JD, health/medicine/public policy reporter (including students or candidates) who tweeted about the risk of Mpox to children and young people in school.

**Exposures:** Tweets containing the keywords ‘school’ and ‘mpox’, ‘pox’, or ‘monkeypox’ from May to October 2022.

**Measures:** The primary outcome was the total of and ratio of accurate vs inaccurate tweets, the latter further subdivided by exaggerating or minimizing risk, and stratified by account author credential type. Secondary outcomes included total likes, retweets and follower counts by accurate vs inaccurate tweets, by month and account credentials. Finally, Twitter user exposure to inaccurate vs accurate Mpox tweets was estimated.

**Results:** 262 tweets were identified. 215/262 (82%) were inaccurate and 215/215 (100%) of these exaggerated risks. 47/262 (18%) tweets were accurate. There were 163 (87%) unique authors of inaccurate tweets and 25 (13%) of accurate tweets. Among health care professionals, 86% (95/111) of tweets were inaccurate. Only health reporters, (23/41) 56% of tweets, were more likely to provide accurate information, however this was driven by one reporter. Multiplying accuracy by followers and retweets, Twitter users were approximately 974x more likely to encounter inaccurate than accurate information.

**Conclusion:** Credentialed Twitter users were 4.6 times more likely to tweet inaccurate than accurate messages. We also demonstrated how incorrect tweets can be quickly amplified by retweets and popular accounts. In the case of Mpox in children and young people, incorrect information exaggerated the risks 100% of the time.

**Key Points:** *Question:* Were tweets during the summer/ fall of 2022 that discussed the risk of Mpox to children and young people in school more likely to be inaccurate or accurate?

*Findings:* Credentialed Twitter users discussing the risk of Mpox to children and young people in schools were 4.6 times more likely to tweet inaccurate than accurate messages. 215/262 (82%) tweets were inaccurate and 215/215 (100%) of these exaggerated risks. 47/262 (18%) were accurate.

*Meaning:* Twitter users were more often exposed to inaccurate than accurate information about the risk of Mpox to children and young people in school.

## Introduction

On May 17^th^, 2022, the first case of Mpox was documented in the United States^1^. Mpox is a DNA virus of the Orthopoxvirus genus of the family Poxviridae which typically causes illness that starts with a febrile prodrome followed by an eruptive phase with a defining rash^2^. Cases grew during the summer months and peaked on August 1^st^ at 645^3^ (Fig. S1). On August 4^th^, U.S. Department of Health and Humans Services Secretary Xavier Becerra announced that Mpox was a Public Health Emergency^4^. By mid-June 2022, data indicated ≥ 95% of cases were in males and 90-99% of cases were men who have sex with men (MSM) and studies continued to show this pattern through July^5,6,7,8,9^ (Fig. 1). Over the summer of 2022, Twitter was utilized by the lay public and health experts alike to provide information, draw awareness to, and make predictions regarding the spread of the Mpox virus.

**Figure 1.**
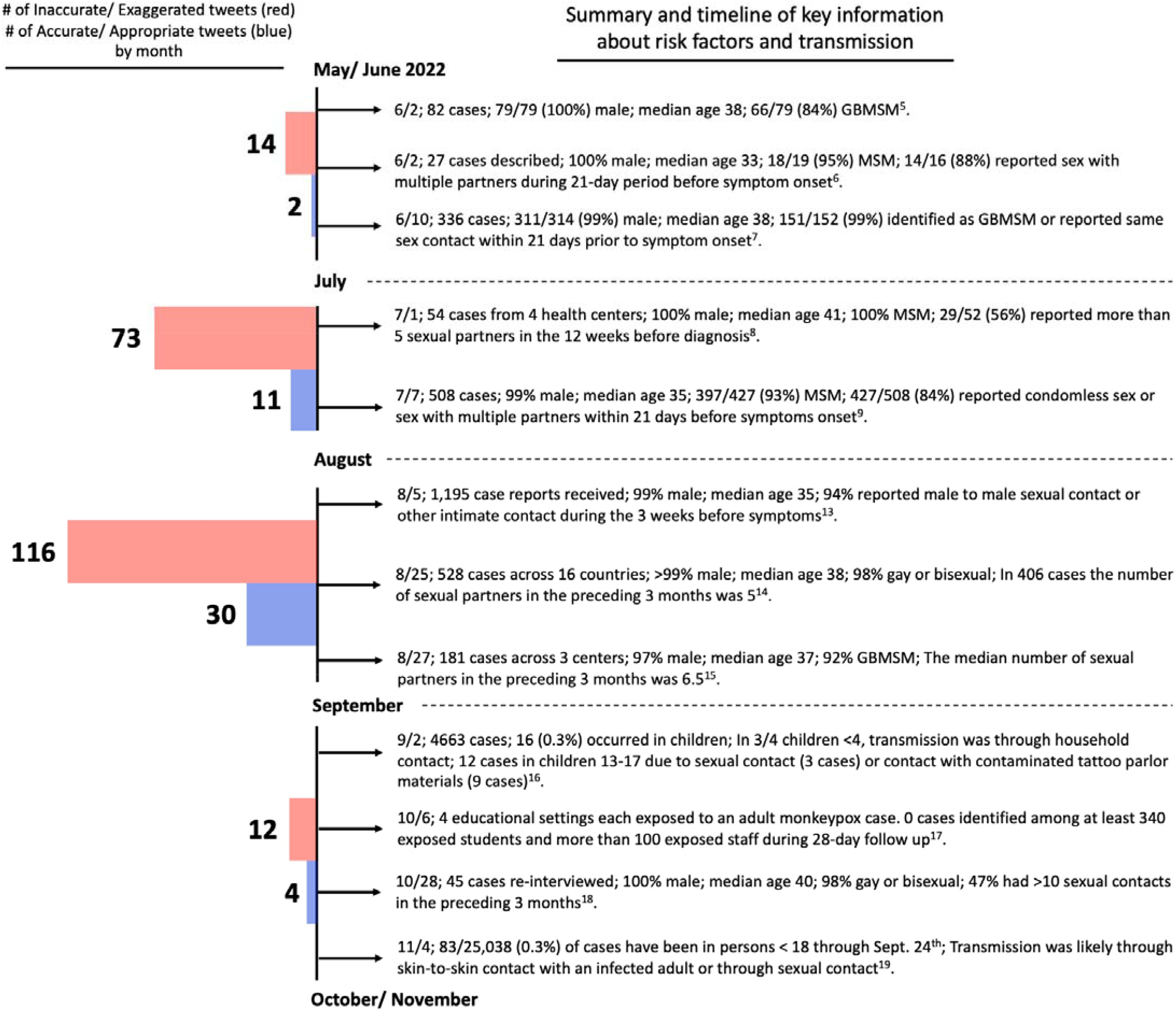
Timeline of risk factors and transmission of Mpox overlayed with the number of inaccurate and accurate tweets per month. Number of tweets that were inaccurate vs. accurate, broken down by month (left). Red bars are inaccurate (all exaggerated) tweets and blue bars are appropriate tweets. Timeline (right) organized by publication date of studies meeting our search criteria, which describe patient characteristics/risk factors in confirmed Mpox cases and transmission. GBMSM = gay, bisexual, men who have sex with men; MSM = men who have sex with men.

Twitter is a social media platform with around 450 million active users of diverse backgrounds, areas of expertise and follower counts. Different emotions may be used by twitter users, and prior work has shown that outrage is a common emotion and that future episodes of outrage may be amplified by positive feedback among other users^10^. One analysis during the COVID-19 pandemic found fear was the most commonly expressed emotion, with around a fifth of all COVID-19 related tweets expressing fear^11^. It has not been investigated how many of those tweets expressed an accurate and appropriate level of fear. One recent analysis of TikTok videos about Mpox found the information in general to be “poor” and “incomplete”, however the extent to which the information was inaccurate or exaggerated was not described^12^. For infectious diseases in general it is unclear how often accounts that are viewed as credentialed experts tweet accurate vs inaccurate information, or to what extent the inaccurate information overstates or understates risks.

We sought to determine how often Twitter users, who might be perceived as domain experts on MPox, public health or infectious disease, provided accurate vs. inaccurate information on risks. We focused on comments regarding schools for children and young people because Mpox was shown early on to be spreading in MSM communities and not among children. Moreover, discussion of this pathogen was used in debates regarding fall school precautions or restrictions for children.

## Methods

### Overview

We created a list of tweets provided by accounts that could be viewed as having specific expertise in Mpox, science, medicine, or public health policy. We sought to describe the ratio of tweets which provided inaccurate to accurate information. We also performed a retrospective literature search to identify studies that described characteristics of patients with Mpox infections. Finally, we determined how many tweets that initially met our search criteria were subsequently deleted. We calculated the ratio of false information (tweets) in the direction of exaggerating risks to children or young people with those which accurately described risks or areas of uncertainty without creating undue fear and ended up being objectively accurate or were objectively accurate at the time.

### Search Strategy

We performed a Twitter advanced search in January of 2023 using the following search terms: school (pox OR monkeypox OR mpox) min_faves:1 lang:en until:2022-10-01 -filter:replies, which would include any tweets meeting our search criteria occurring prior to October 1st, 2022. The term “pox” was used in addition to “monkeypox” as many users referred to monkeypox as “monkey pox”. To ensure that tweets occurring before the Mpox outbreak in the U.S. were captured, the search strategy did not include a start date. We stopped our search October 1st, corresponding with a decline in the number of Mpox cases (Fig. S1)^1^. The search strategy was adapted to exclude tweets that were replies (comments) because replies receive less engagement and are overall, less impactful. Additionally, we utilized the snowball method to retrieve additional tweets or replies that fit our inclusion criteria but did not appear in the Twitter advanced search. In April 2023, we re-reviewed all the included tweets to determine how many and which percent had been deleted.

### Inclusion Criteria for Tweets

One author (BK) analyzed all tweets in the Twitter advanced search. First, the author and biography of each tweet was viewed and immediately excluded unless at least one of the following set of credentials was met: MD, DO, nurse, pharmacist, physical therapist, other health care provider, PhD, MPH, other Ed. degree, JD, health/medicine reporter/journalist/columnist/expert, public policy reporter (students or candidates of these professions were also included). We categorized MDs, DOs, nurses, pharmacists, physical therapists, others who provide direct patient care, and students in these degree programs as “health care”. PhDs and MPHs and other Ed. degrees as well as students in these degree programs were placed in a second category. JDs were in their own category and all science/medicine journalists were placed in a fourth category called “health reporters”.

All tweets meeting inclusion criteria by author one (BK) were then reviewed for appropriateness for inclusion by a second author (TBH). All included tweets were subsequently analyzed for content by the same two authors (BK, TBH). Tweets were placed into two categories: tweets that were inaccurate/exaggerated or understated risks and tweets that were accurate/appropriate.

Inclusion criteria for inaccurate tweets (either exaggerated or minimized risk) included: overstating the risk of Mpox infection in children, predicting that Mpox would spread widely in schools, recommending schools adopt mitigation measures (e.g. masking, vaccines, etc.) to prevent Mpox spread, that schools should be closed or delayed to prevent or delay Mpox transmission, recommending vaccinating very low risk groups for Mpox infection, messaging that provokes fear without supporting evidence, understating risk of behaviors repeatedly shown to be high risk for transmission.

Inclusion criteria for accurate tweets (appropriate or non-misleading) included: reporting information in accordance with prevailing evidence – the evidence shows that MSM individuals are at highest risk of infection, reassuring messaging that schools are not a high-risk environment for transmission and that children are not a high-risk population for infection, messaging that is balanced, sensible, and follows the current data, provides neutral information which is not factually incorrect.

### Mpox Literature Search

At the time of our analysis, there had been multiple studies characterizing risk factors and transmission dynamics of Mpox. We search PubMed using the following search terms: “Monkeypox virus”[Mesh] OR “Monkeypox”[Mesh] with the Human and 2022 filter. We selected publications from this list that described cases of Mpox and risk factors for transmission.

Data published between June and October 2022 showed that Mpox infection was predominantly in males between the ages of 30-40 who identify as MSM^5,6,7,8,9,13,14,15,18^. Additionally, a significant number of Mpox infections were in individuals engaging in sex with multiple partners or those who have a pre-existing immunocompromising condition. We did not identify any studies showing that schools are a high-risk environment for transmission or that school children are a high-risk population of contracting the virus^16,17,19^. We used the data provided in these studies to define our framework for determining if a tweet was “inaccurate/ exaggerated risks” vs “accurate/ appropriate”.

### Data Extraction

In January 2023, we extracted the date of the tweet, tweet text, credentials of the author, number of favorites, number of retweets, and number of followers of the tweet’s author. In April 2023 one author (BK), reviewed all tweets through the Twitter platform to determine if tweets were still posted or if they had been deleted by the author.

### Data Analysis

We calculated a ratio of inaccurate to accurate tweets and specified whether the inaccurate tweets exaggerated or minimized risks. Additionally, we separated all tweets into 4 bins based on the credentials of the author: health care (MD, DO, nurse, pharmacist, physical therapist, other health care practitioner, students); health/ medicine reporter/ journalist/ columnist/ expert; PhD, MPH, other Ed. degree (students included); JD (students included).

We calculated the ratio of inaccurate/ exaggerated to accurate/ appropriate tweets within each bin. We calculated the same ratio based on the month the tweet was published. We also compared the mean and median number of account followers between tweets which were inaccurate and accurate as well as mean and median number of likes and retweets for each tweet. Finally we used the Wilcoxon Rank-sum test in R (version 4.2.2) to compare the distribution of the median follower count for tweets that were inaccurate/ exaggerated to those providing accurate/ appropriate information. R was also used to generate the waterfall plot showing the number of tweets per user and to create a Kaplan-Meier plot of the cumulative number of likes and retweets over time. The Wilcoxon Rank-sum test in R was used to compare the cumulative number of likes and retweets for inaccurate/ exaggerated and accurate/ appropriate tweets. Excel was used to tabulate descriptive statistics. In accordance with 45 CFR §46.102(f), this study was not submitted for institutional review board approval because it involved publicly available data and did not involve individual patient data.

## Results

We identified 262 tweets spanning May 18th, 2022, to September 19th, 2022. The 262 tweets came from 188 unique accounts. Among these unique accounts, 187 were individuals and 1 was an institution. Among individuals, 48% (90/187) were categorized as health care, 7%, (13/187) were health reporters, 39% (72/187) were PhD, MPH, or other Ed. degree, and 6% (12/187) were JD. The median tweets per account was 1 (IQR 1 to 1).

215 (82%) tweets overstated or exaggerated the risk of Mpox to children or young people in the school setting, while 47 (18%) tweets provided accurate or appropriate information. The ratio of exaggerating risks to accurately stating risks was 4.6:1. Ten selected examples of inaccurate/ exaggerated tweets are in **Table 1**, and the full dataset is in the supplement.

**Table 1.**
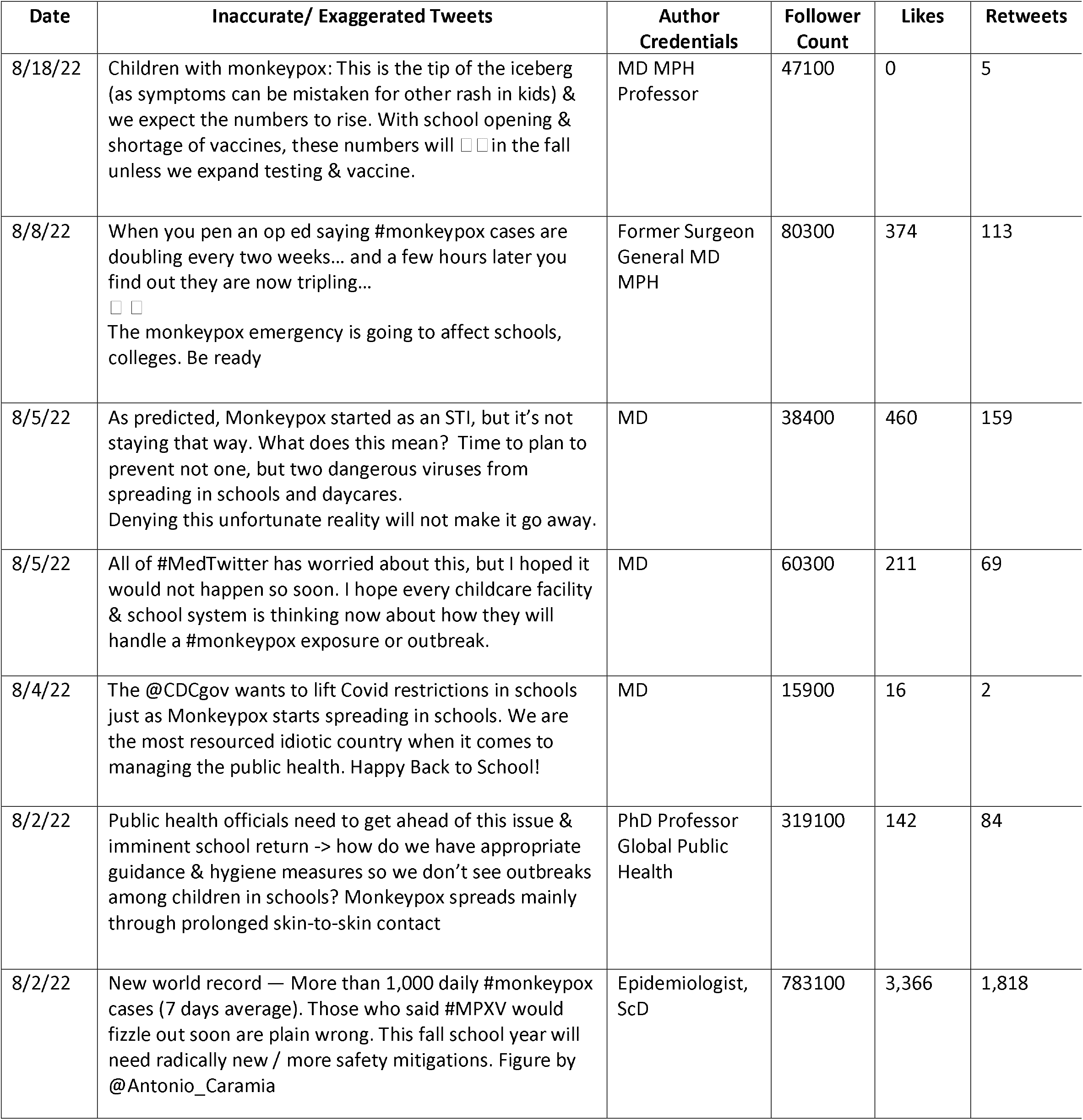
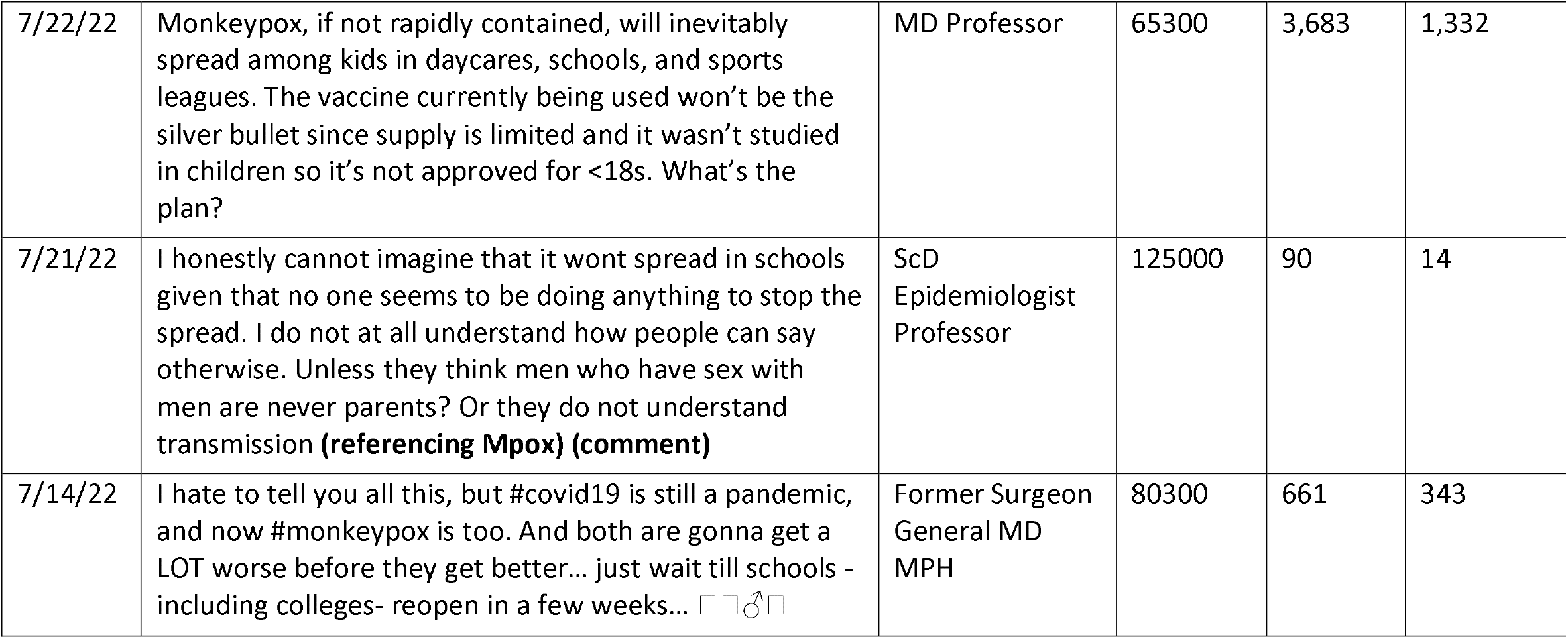
Selected examples of inaccurate/ exaggerated tweets. Bolded text in the Tweet column is notes inserted by investigators. Tweets that are comments are denoted with a bold (comment). Tweets that were deleted are denoted with a bold (deleted tweet).

The ratio of inaccurate/ exaggerated to accurate/ appropriate tweets in May/June, July, August, and September was 7:1, 6.6:1, 3.9:1, and 3:1 respectively (**Fig. 1**). August had the highest number of tweets from both categories: 116 tweets that were inaccurate and 30 that were accurate.

The percentage of tweets that were inaccurate/ exaggerated by the credentials of the author were: Among health care professionals 95/111 (86%), health reporters 18/41 (44%), PhDs/MPH/Other Ed. 93/97 (96%), JDs 9/13 (69%) (**Fig. 2**). Only health reporters were more likely to provide accurate information, though this is based predominantly on 1 outlier (**Fig. 3, rightmost bar**).

**Figure 2.**
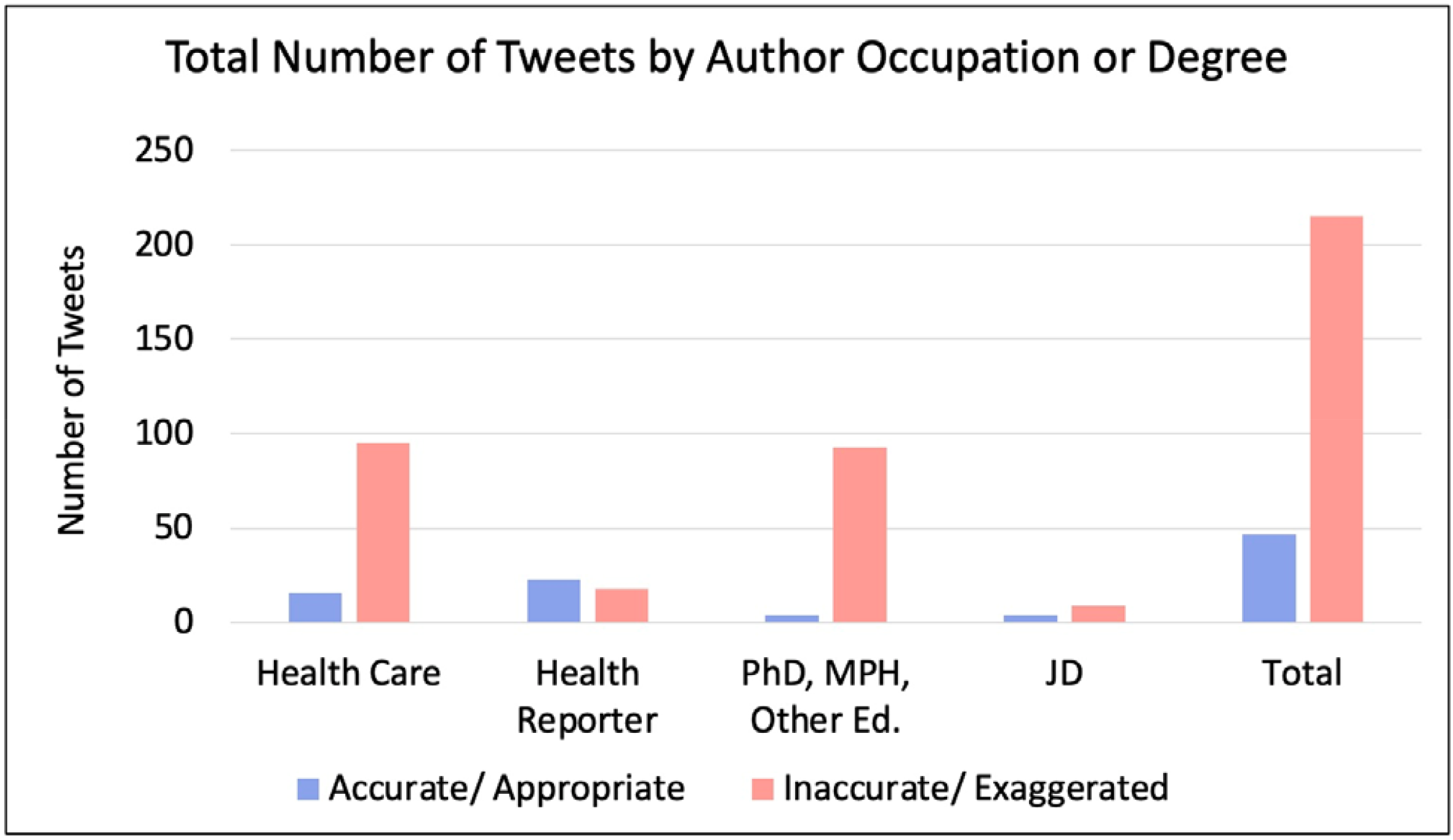
Inaccurate and accurate tweets by author occupation or degree. The number of tweets categorized by the author’s occupation or degree and whether the tweet was inaccurate or accurate.

**Figure 3.**
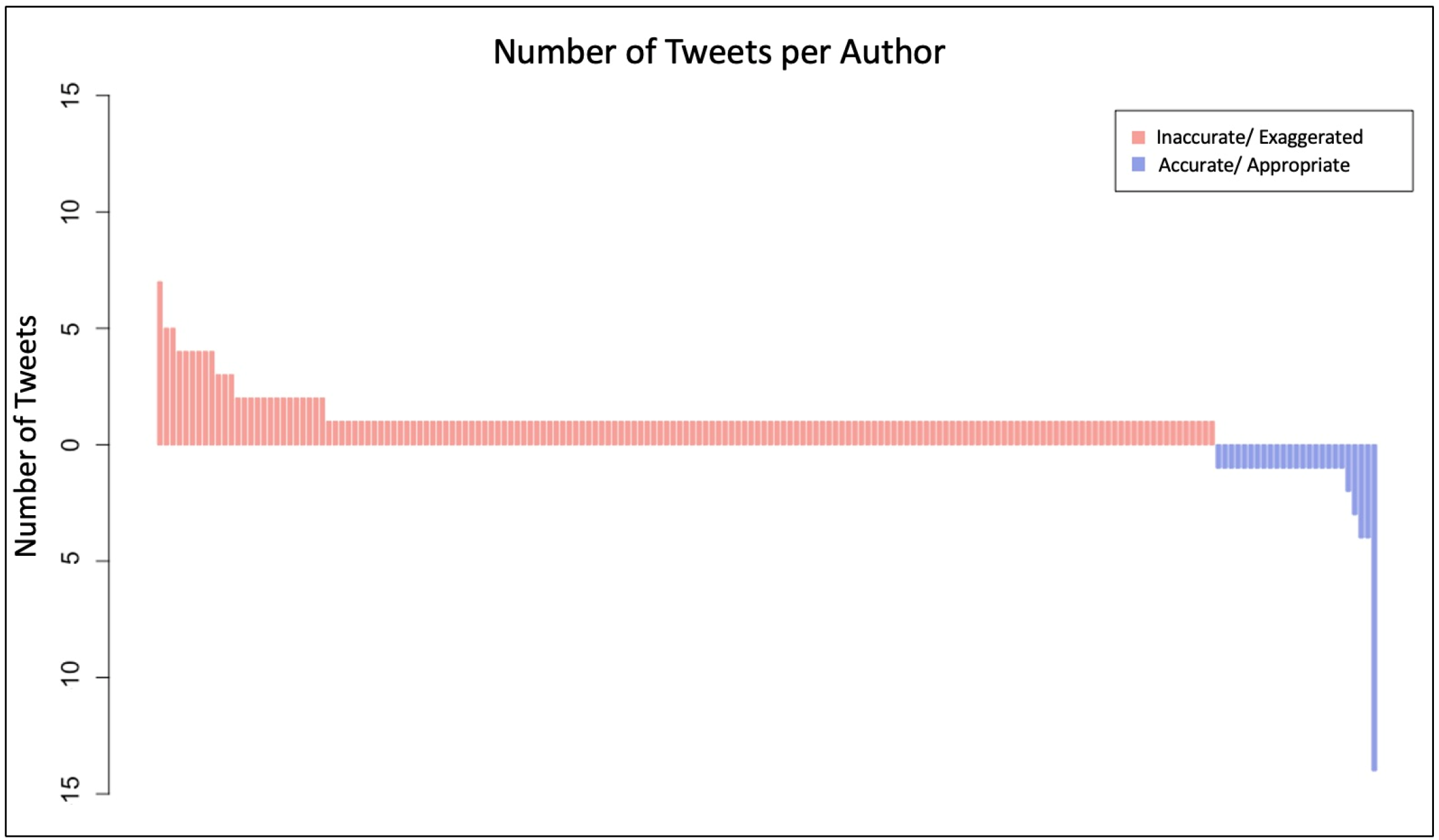
Number of tweets per author. Waterfall plot showing the number of tweets per author. Each bar represents a unique author. The red bars represent authors who had inaccurate tweets and the blue bars indicate authors who had accurate tweets.

There were 163 unique authors of the 215 tweets that overstated the risk (**Fig. 3**). There were 12 individuals with at least 3 tweets within this group. In contrast, we found 25 unique authors of the 47 tweets that provided accurate information (**Fig. 3**). There was one author in the Health Reporter group who had 14 accurate tweets.

The mean and median follower count of the users with exaggerated tweets was 37,229 and 6,409 (Q1 2,658 – Q3 19,900) respectively. For accounts that provided accurate information the mean and median follower count was 31,334 and 17,700 (Q1 10,100 – Q3 17,700). The distribution of the median follower count between users that exaggerated risks and those providing accurate information was statistically significant (p = 0.00014).

The cumulative number of likes over all tweets was 201,811 and 7,084 (28.5-fold difference) for those that were inaccurate and accurate, respectively (p<0.001) (**Fig. 4a**). However, there were two inaccurate tweets that were outliers and had a significant number of likes: 126,000 and 41,600. The cumulative number of retweets over all tweets was 50,710 and 1,295 (39.2-fold difference) for those that were inaccurate and accurate, respectively (p<0.001) (**Fig. 4b**). The same two tweets were outliers with 33,502 and 6,477 retweets. The cumulative number of account followers was 8,004,244 and 1,472,683 for inaccurate and accurate tweets, respectively.

**Figure 4.**
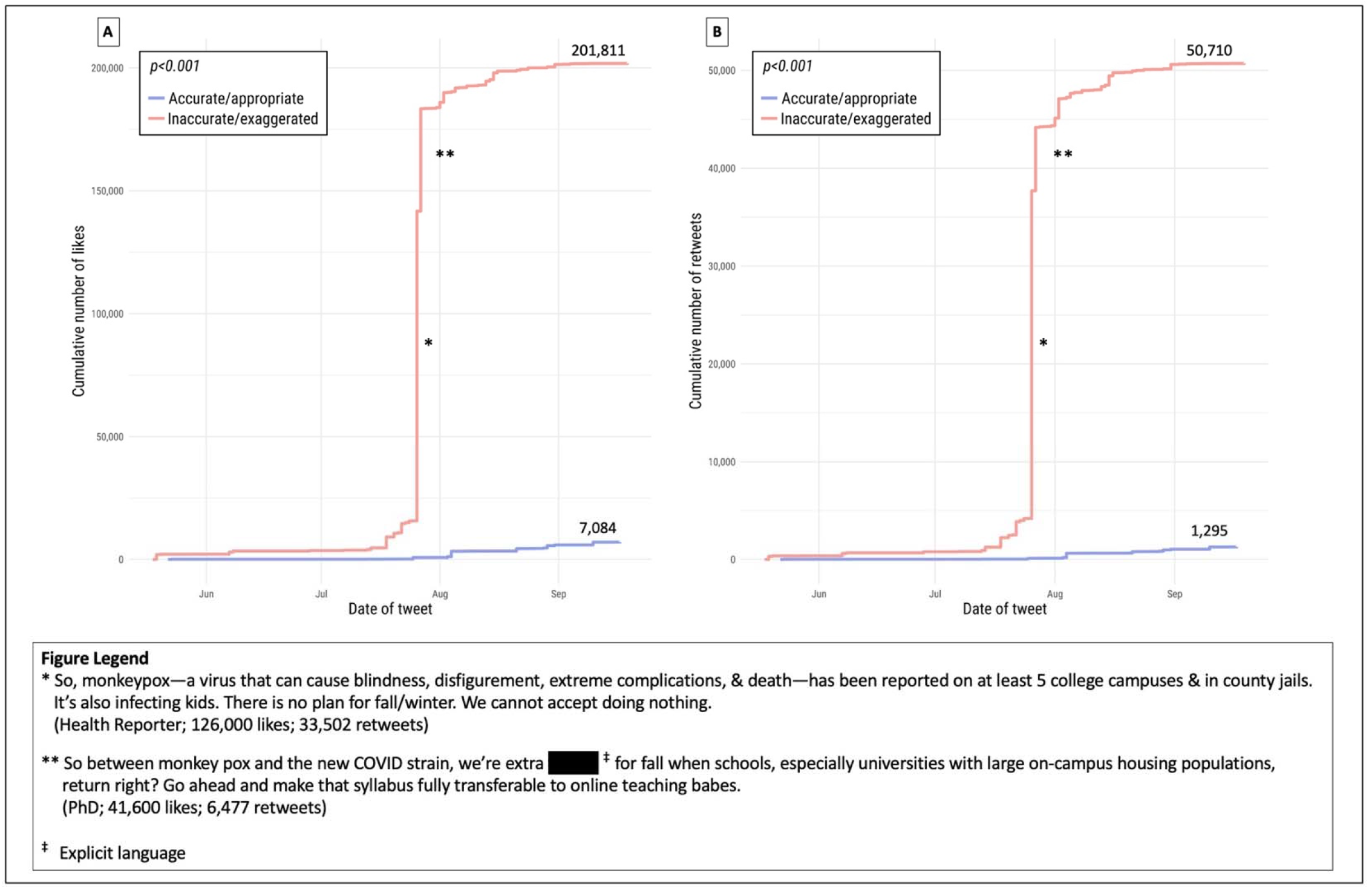
Cumulative number of likes and retweets over time. Kaplan-Meier plot showing the cumulative number of likes **(A)** and retweets **(B)** over time grouped by either being inaccurate or accurate. The figure legend denotes the two tweets that contributed substantially to the total sum of likes and retweets.

There were 9 tweets that appeared to be deleted at a point between our initial search and April 2023. All tweets were in the exaggerated category. There were 4 tweets in the accurate bin and 6 tweets in the inaccurate bin that were comments/replies. These were retrieved through the snowball method.

## Discussion

Our study found health care professionals and other credentialed Twitter profiles tended to provide inaccurate information, which exaggerated the risks of Mpox in the school setting. Tweets exaggerating risks exceeded accurate ones by a ratio of 4.6:1. The profession/degree category of “PhDs, MPHs and other Ed. degrees” had the highest tendency to overstate risks at a ratio of 23:1. Health reporters were the only category which was slightly more likely to tweet accurate vs inaccurate and fear-based information by a ratio of 1.3:1. However, this was driven by one reporter who was an outlier tweeting 14 accurate tweets about Mpox included in our study. Health care providers also performed very poorly in terms of accuracy and their tendency to exaggerate risks was 5.9:1.

Twitter is a free and readily accessible site with an obvious potential to educate and inform the public. However, our study highlights potential drawbacks of relying on Twitter profiles, even credentialed accounts, for health care information. While we are unable to determine the reasons the information provided about Mpox risk to children and young people in the school setting tended to overstate risks, it is noteworthy that the accounts that *did* exaggerate had on average higher follower counts, likes and retweets to the individual tweets.

Tweets that were inaccurate came from accounts that on average had 19% higher follower counts and the individual inaccurate and exaggerated tweets were liked and shared 28.5× and 39.2× more frequently than accurate tweets, respectively. Thus looking at the ratio of inaccurate to accurate tweets underestimates the actual impact the inaccurate tweets had vs those that were found to be accurate.

We do not have view counts for each tweet (as these were recently and not retrospectively added to Twitter), but multiplying the ratio of inaccurate to accurate by a share differential for inaccurate vs accurate tweets by the follower number and retweet difference for each tweet suggests the actual number of views could be estimated at (4.6 × 5.4 × 39.2) would give a 974 fold higher likelihood on average of seeing inaccurate (and exaggerated) vs accurate information about Mpox in children and young people in the school setting. Our study demonstrates the potential of Twitter to magnify inaccurate and fear-based messaging. In fact the 974-fold difference is likely to be an underestimate considering the inaccurate tweets may have been more likely to be deleted by the author before we had identified them in our search.

August had the highest number of inaccurate/ exaggerated tweets. It is important to note that at this time there had already been a number of published studies showing that high risk individuals were those who identify as MSM and that sexual activities were the highest risk behaviors for infection (**Fig. 1**). Although Mpox can spread through routine skin-skin contact, there was no evidence to suggest that schools would be a favorable environment for viral transmission. Thus, it is puzzling why so many perceived health experts arrived at the conclusion that school would be dangerous for children and young people, that schools must adopt mitigation measures to prevent viral spread, and that in extreme cases, schools should be delayed or closed during the outbreak. It may be likely that August had the highest number of tweets because of the temporal relationship with the start of the U.S. school year.

One user who tweeted multiple inaccurate/ exaggerated tweets offered a justification for the content of the original tweets:

*Did you stop to think maybe raising the alarm did what it was supposed to do? Ie encouraged the administration to massively increase availability of vaccinations and testing, and helped prevent more spread (again except in black and brown communities…). I’m glad for progress!*^*20*^

Although increased availability of vaccinations and testing may rightly be considered a net positive, using this to defend messaging that is inaccurate and exaggerated is problematic because it neglects to acknowledge or even recognize the harms associated with the narrative. Parents may have suffered anxiety and stress about the health and safety of their children in school. In addition, children and young people may have been worried or hesitant about attending school and playing with friends.

The field of public health and more generally medicine, relies on the public’s trust, but trust is difficult to obtain and may be even more challenging to keep. Public messaging around monkeypox raises the concern that some trust was forfeited due to inaccurate messaging.

Unlike in real-life medical and health settings, it is unclear if accounts that tweet inaccurate or fear-based information face any repercussions if the advice or information they provide is incorrect. In fact, our study suggests that tweets that were incorrect tended to come from accounts that on average had a higher number of followers. Twitter users which tweeted inaccurate information also have the option of going back and deleting earlier tweets which were incorrect (as we saw in 9/215 inaccurate tweets), though the advice had already been disseminated months earlier at that point. It is also not clear how many inaccurate tweets had already been deleted by the time we did our initial search in January of 2023, several months after the Mpox cases started steadily declining.

### Strengths and limitations

Our study has 3 strengths and 4 limitations. To our knowledge, ours is the first to judge accuracy of public health information on Twitter for an emerging infectious disease. We provide specific information on whether accurate or inaccurate tweets overstated or understated risks by profession/degree and follower counts, likes and retweets. We demonstrate that inaccurate tweets are able to reach far more readers than accurate ones, supporting the adage, ‘a lie will fly around the whole world while the truth is getting its boots on’. An added strength of our work is we quantify erroneous tweets in the context of a vulnerable group (children and young people) which may be viewed as more vulnerable to the impacts of inaccurate health information and overstated risks.

Regarding limitations, our criteria for accurate vs inaccurate tweets were based on a literature review and it is natural that others will feel differently. However the included tweets were reviewed for accuracy independently by two authors and we have made all the tweets available for reanalysis in the supplementary material and encourage re-analysis. Second, the accuracy of the Twitter Advanced Search function has not been described in the scientific literature or made publicly available in any way that we are aware of. The search function may have biased results by removing or censoring certain tweets. Or it may have missed relevant tweets. Specifically, it is possible some users deleted tweets in retrospect viewed to be embarrassing or incorrect. Third, ours is an investigation of one important infectious disease in 2022 and may not apply to other contexts. Finally, we were unable to state what the impacts of the incorrect and exaggerated information may have been on children, their schools and society in general, though sound decisions are seldom based on false premises.

## Conclusion

Our study found that credentialed Twitter users were 4.6 times more likely to tweet inaccurate vs accurate information about Mpox risks in the school setting in children and young people. 100% of the inaccurate information exaggerated the risks of Mpox in this population. These accounts had more followers on average and higher cumulative likes and retweets than accounts that tweeted accurate information.

This finding may have major and widespread societal ramifications attributable to inappropriately heightened anxiety, and subsequent loss of trust. Those seeking health information from Twitter should be aware of our documented high rates of inaccuracy even from the accounts of credentialed health professionals.

## Supporting information

Supplemental Tables and Graphs

## Data Availability

All analyzed tweets are located in the supplement.

