## Supplemental Tables and Graphs for "Analysis of tweets discussing the risk of Mpox among children and young people in school (May-Oct 2022): Public health experts on Twitter consistently exaggerated risks and infrequently reported accurate information"

**Supplementary Appendix**

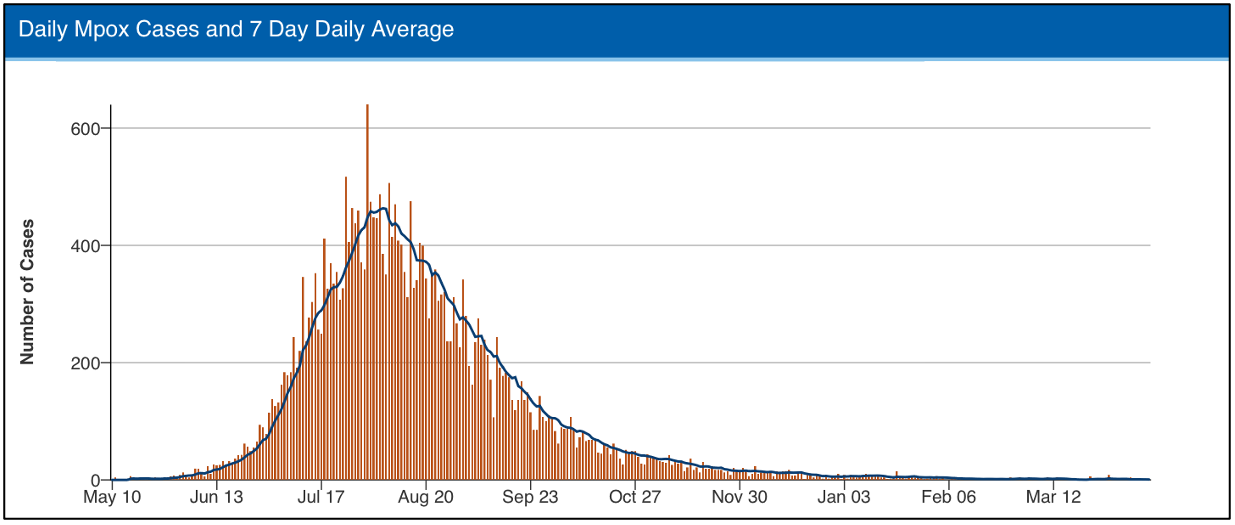

**Figure S1.** CDC data showing the daily Mpox case count (each red bar represents one day) and the 7 day daily average number of Mpox cases (dark blue line).

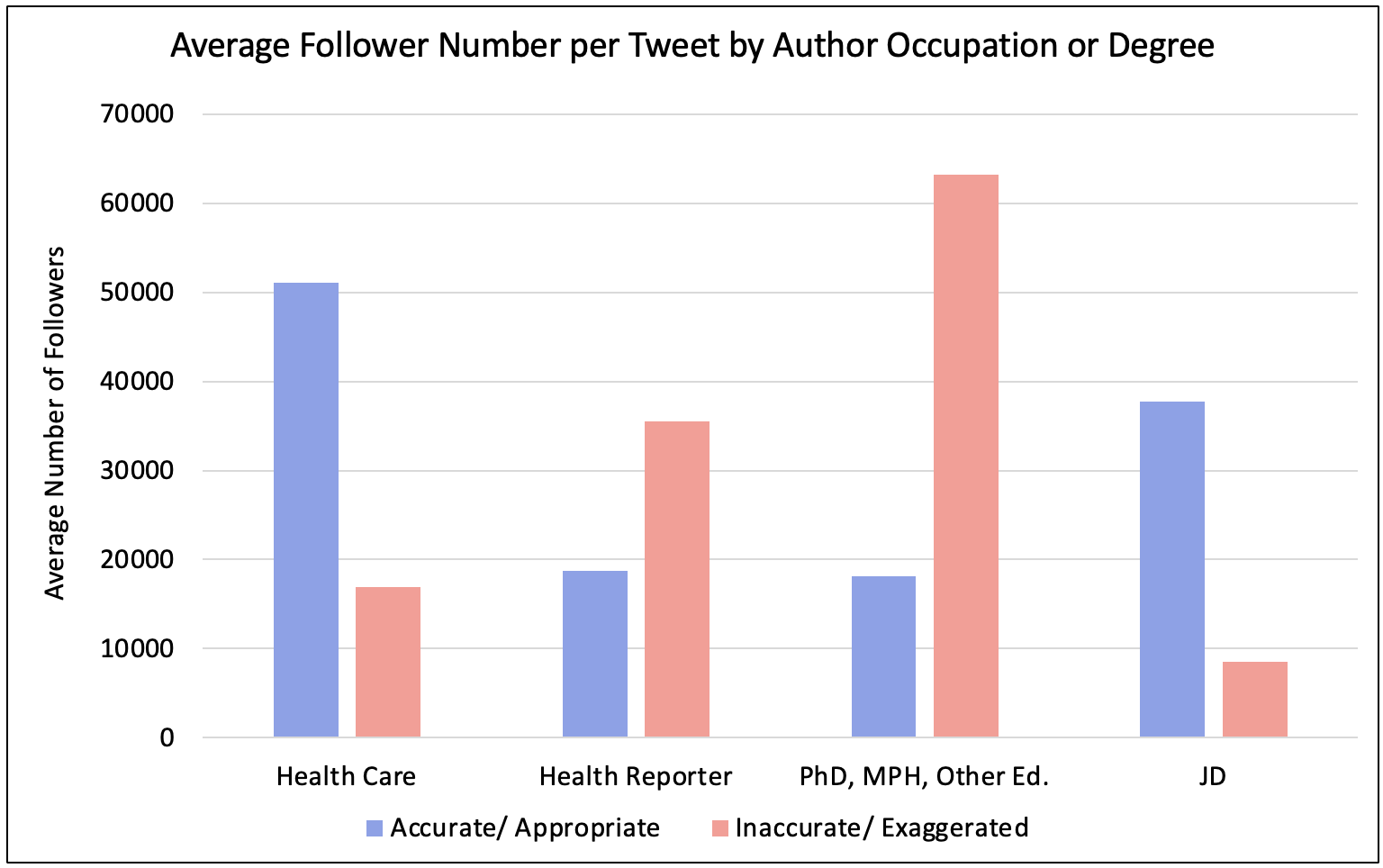

**Figure S2.** Average follower number per tweet categorized by author occupation or degree. All tweets were considered independent even if one author had multiple tweets.

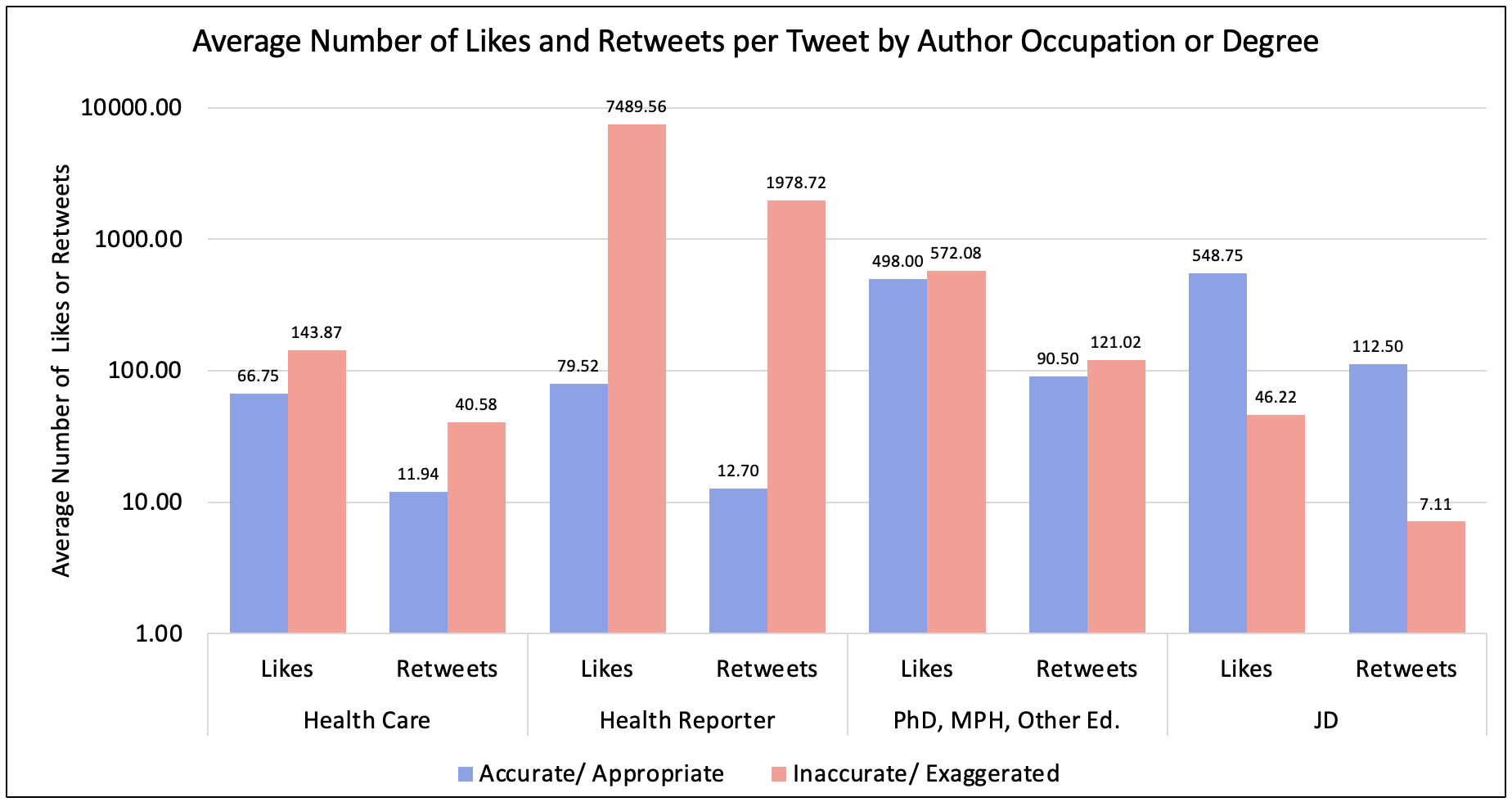

**Figure S3.** Average number of likes and retweets per tweet categorized by author occupation or degree. The graph is log scale.

**Table S1.** Average number of followers, likes, and retweets per tweet categorized as either accurate/ appropriate or inaccurate/ exaggerated and by author occupation or degree. The Q1 to Q3 range is also provided. There were two tweets in the inaccurate/ exaggerated category that were missing data for likes and retweets.

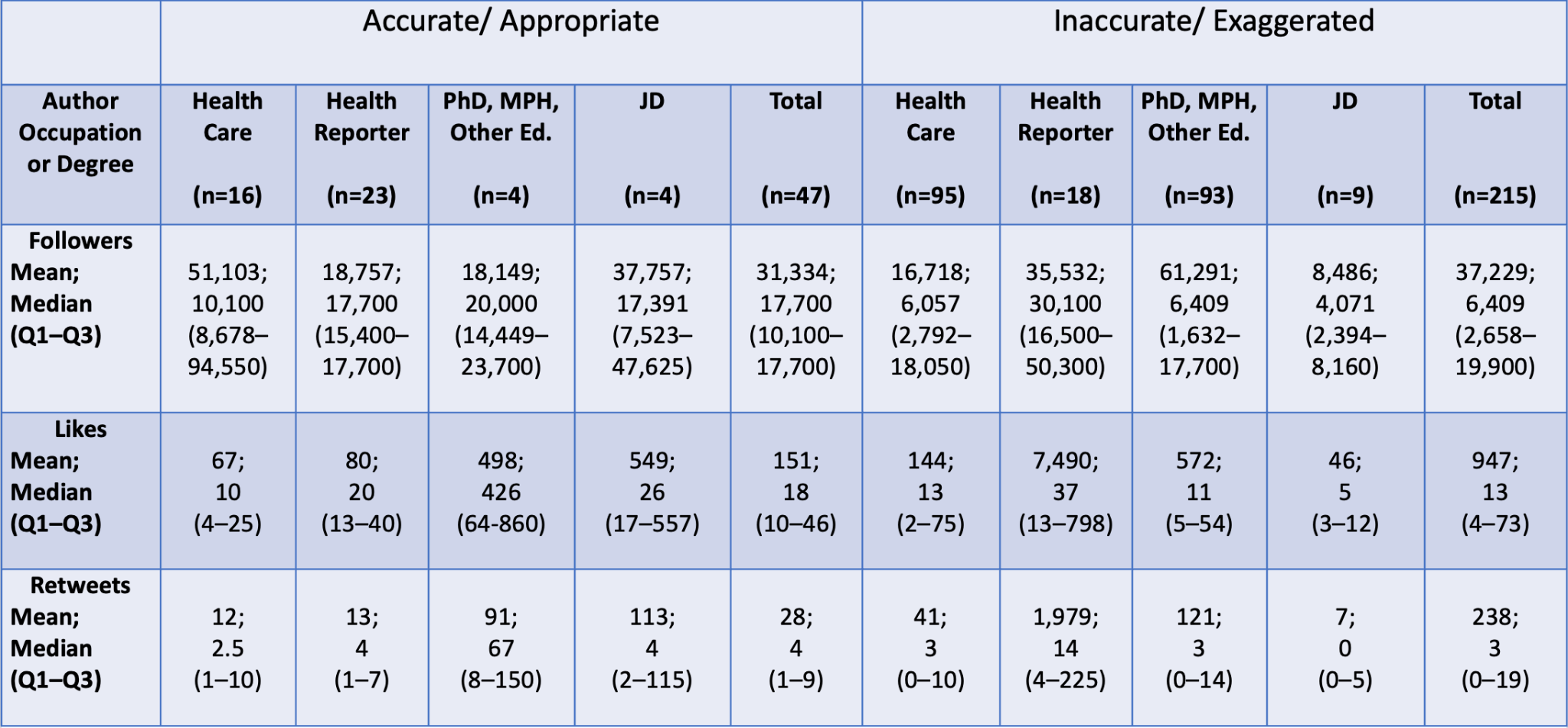

**Table S2.** All tweets categorized as accurate/appropriate. Bolded text in the Tweet column is added notes by the investigators. Tweets that are comments are denoted with a bold (comment).

| **Health Care** |  |  |  |  |  |
| --- | --- | --- | --- | --- | --- |
| **Date** | **Tweet** | **Author Credentials** | **Follower Count** | **Likes** | **Retweets** |
| 9/17/22 | MONKEYPOX TRANSMISSION: Now 4 months into outbreak (1st case reported May 12) & overwhelming majority of cases in gay men. 2 concerns abating CDC shows no transmission to health care workers- clinic https://cdc.gov/mmwr/volumes/71/wr/mm7138e2.htm?s_cid=mm7138e2_x… WHO -no school transmissions https://worldhealthorg.shinyapps.io/mpx_global/ | MD MPH Professor | 94000 | 24 | 9 |
| 9/9/22 | COVID, monkeypox, and polio are of concern for parents as they send their children back to school. But what actually poses risk? I joined  @KoriOnAirand @ArrianeeLeBeau on New York City's  @PIX11News to explain. **(video explaining how risk of Mpox transmission is low)** | MD | 14500 | 1 | 1 |
| 8/29/22 | 1/ Some parents are worried their kids might be at risk for monkeypox at school. How worried should they be? https://cbsnews.com/video/more-monkeypox-vaccines-to-be-distributed-us/#x… on  @CBSNews with @TanyaRivero &  @tonydokoupil **(video explaining that infection is rare in children and most transmission between children happens sexually)** | MD ScM Professor | 115700 | 9 | 4 |
| 8/26/22 | How to Protect Against Monkeypox as School Starts Experts say children are not at a high risk of infection. But they have advice to keep everyone — from toddlers to college kids — safe. https://nytimes.com/2022/08/17/well/monkeypox-kids-school-day-care.html… #student #school #health #prevention #empathy #BetterTogether | MD Professor | 4804 | 1 | 1 |
| 8/25/22 | School's back. Parents have Qs about #monkeypox.  Can it be spread by being next to someone in a classroom or playground? Are there certain activities for K-12 students that are higher risk? What about for college students? With @katiahetter @CNN: **(links to an article explaining how the risk of Mpox spreading to children through everyday contact is rare)** | MD Professor | 178000 | 11 | 3 |
| 8/23/22 | These are articles from  @MedStarHealth that everyone should read, especially all the Mopoxians, who are already trying to close schools & reinstate mask mandates. MONKEYPOX IS NOT THE NEXT PANDEMIC! Read that again. And again… https://app.care.medstarhealth.org/e/es?s=145898957&e=20719&elqTrackId=55395a9adee64c6688c9f33ff191da8d&elq=6af2f798e79143cb99bcfcb0e020b41a&elqaid=910&elqat=1… https://medstarhealth.org/blog/monkeypox-outbreak | Mental Health Therapist | 584 | 2 | 0 |
| 8/14/22 | If you are worried about #monkeypox and kids as they return to school in the next few weeks, then this short article is worth reading | MD Professor | 10100 | 13 | 1 |
| 8/12/22 | If my kid comes back from school with monkeypox someone is gonna get their heads beat in | MD | 9969 | 4 | 0 |
| 8/10/22 | I’ve heard parents are worried about things that are good for kids & families—like swimming lessons or school. I get it, given misinformation about monkeypox. It’s so reassuring to hear from  @Larry_Kociolek  that the risks are very low & we should keep doing the things we value. | MD Professor | 627 | 6 | 2 |
| 8/3/22 | This is unnecessary scaremongering for parents. In England, several 100 students were exposed to an adult with #monkeypox in school before summer holidays (when there were no mitigations) & there were ZERO secondary cases in the students - their risk in school is less than 0.5% | MD Professor | 10100 | 326 | 64 |
| 7/27/22 | Monkeypox cases are ⬆️ globally but kids continue to have very low risk. In the UK, there has been only 1 child case so far, even though there have multiple occasions where kids came into contact with an adult with monkeypox disease at home or in school | MD Professor | 10100 | 12 | 5 |
| 7/25/22 | Let me be very clear, there is no data to show masking kids, closing schools, and zoom schooling is effective against #monkeypox | RN | 96200 | 3 | 0 |
| 7/25/22 | Reading what people are saying about monkeypox and schools on here is horrible. Who are these people? Germophobics with long lockdown anxiety or bots? | MD | 213700 | 533 | 63 |
| 7/14/22 | Parents in northumberland might also want to read this. Remember risk of catching monkey pox at home/ school is VERY low ( less than 0.5%) thanks @ShamezLadhani #FactsMatter  @StamfordhamPS @Darrashallprim @GreatNorthCH @PontHigh @primarypont @throckleyprim @HeddonSchool | MD | 1257 | 7 | 2 |
| 7/13/22 | 1/ Parents in London may be concerned about the recent media report of a reception class being sent home after being exposed to a #monkeypox case The risk to kids is very low but we are being cautious until we are certain - here’s what we know …🧵 | MD Professor | 10100 | 28 | 14 |
| 5/23/22 | This guy is just the worst.   It’s completely unhelpful & inaccurate fear-mongering to describe #monkeypox as “surging” or “out of control.”  Reminder that a PhD in nutrition & dropping out of medical school does not remotely make him an expert in poxviruses. | MD | 47900 | 88 | 22 |
| **Health Reporter** |  |  |  |  |  |
| **Date** | **Tweet** | **Author Credentials** | **Follower Count** | **Likes** | **Retweets** |
| 9/15/22 | CDC director Rochelle Walensky reported no evidence of substantial spread of #monkeypox outside of men who have sex with men. There has been no substantial spread in schools or universities. If there are cases there, Walensky said, they see "terminal chains" of infection. **(comment)** | Health and Science Reporter | 17700 | 18 | 7 |
| 8/31/22 | Eric Feigl-Ding continues to spread hysteria about #monkeypox, jumping to conclusions without knowing the details of these cases in Texans <18 y/o (who might be teens). Meanwhile,  @WHO reports no known cases of viral transmission in schools anywhere in the world.  @DrEricDing | Health and Science Reporter | 17700 | 280 | 72 |
| 8/29/22 | There have been zero documented cases of #monkeypox transmitting in school and no substantial transmission chains among kids’ peer groups,  @WHO reports. Every news story that broaches the subject implies to parents that they actually should fear this outcome. 1/ | Health and Science Reporter | 17700 | 20 | 4 |
| 8/29/22 | Just to be clear, @WHO reports 140 people under age 18 with #monkeypox. @CDCgov reports <20 in US. There has been no known transmission in schools. Official data lags, but cases in children have remained well under 1% for 3 months. 1/ | Medical Journalist | 15200 | 945 | 135 |
| 8/28/22 | The  @WHO has identified no cases of #monkeypox transmitting in schools anywhere in the world. | Health and Science Reporter | 17700 | 179 | 25 |
| 8/26/22 | Great thread on the monkeypox data worldwide. Overwhelmingly, MPX is spreading like an STI. Out of ~22,000 cases, *zero* were from a suspected exposure in schools. | Health, History & Politics Columnist | 68300 | 21 | 4 |
| 8/24/22 | Kids are at low risk for #monkeypox in schools. It's mainly transmitted via sex or other close skin-to-skin contact. Kids masking at school isn't going to make a difference. | Medical Journalist | 15200 | 15 | 0 |
| 8/22/22 | Echoing their general community guidance Last week's  @CDCgov recommendations for monkeypox in schools says that for students or staff exposed to infections "do not need to be excluded from an educational setting in most cases." https://cdc.gov/poxvirus/monkeypox/schools/faq.html | Reporter on Federal Responses to Public Health Issues | 8120 | 8 | 9 |
| 8/21/22 | There is NO evidence of #monkeypox transmission in schools anywhere around the world. <https://worldhealthorg.shinyapps.io/mpx_global/> **(comment)** | Health and Science Reporter | 17700 | 12 | 2 |
| 8/21/22 | The big question is this: If the likelihood that any child will show up at school with #monkeypox is incredibly remote, then why publish a massive article about this chance? The fact that the article exists implies to worried parents that the risk is substantial. **(comment)** | Health and Science Reporter | 17700 | 29 | 3 |
| 8/21/22 | The rest of the  @nytimes  article on the notion that #monkeypox will pop up in schools did a decent job of continuously reminding the reader that cases in kids are rare. The  @WHO  reported this week there has been no evidence of sustained transmission chains in women or children. | Health and Science Reporter | 17700 | 45 | 0 |
| 8/21/22 | This is how #monkeypox really transmits in the overwhelming majority of cases: not through casual contact at school, work, or crowded spaces, but through sex between men. **(comment)** | Health and Science Reporter | 17700 | 47 | 7 |
| 8/20/22 | This is unscientific fear mongering about #monkeypox. There have been no sustained transmission chains of the virus among children, @WHO reported this week. Cases in kids are rare. And it's not as if kids haven't been in school in Australia and in daycare all over this summer. | Health and Science Reporter | 17700 | 26 | 5 |
| 8/17/22 | "Attending school is unlikely to put people at risk of a monkeypox exposure." | Social Policy, Education, Politics Reporter | 15400 | 6 | 0 |
| 8/8/22 | Regarding the theory that once school starts we'll see an explosion of #monkeypox cases in youths:  This presumes that everyone age 5–23 spends the entirety of June through August in isolation.  No summer camp?  No summer school? No daycare? No sleepovers? Seriously? | Health and Science Reporter | 17700 | 49 | 4 |
| 8/3/22 | #Monkeypox is not COVID. The risk of kids getting it is very low. The few kids who got it in this outbreak fully recovered. If a kid has MPX, keep them home. Maybe take measures to reduce skin-to-skin contact among kids. But school closures are overkill & masks are irrelevant. | Medical Journalist | 15200 | 28 | 3 |
| 8/3/22 | Alarmist U.S. reporting about monkeypox. We should keep our schools clean and provide paid leave so parents can care for sick children at home. Sensible, simple stuff that works for all diseases. | Social Policy, Education, Politics Reporter | 15400 | 13 | 0 |
| 7/30/22 | If you doubt that the #monkeypox outbreak is truly driven overwhelmingly by sex between men and that there is somehow a shadow epidemic in women and kids or there will be an explosion of rampant spread in schools or via toilets, check out all this evidence to the contrary: 🧵⬇️ | Health and Science Reporter | 17700 | 13 | 4 |
| 7/27/22 | We have some major school hesitancy issues right now in the United States and districts like  @apsupdate offering totally unacceptable remote options. I hope that experts like @CarlosdelRio7will be careful when talking about monkeypox and children.  https://ajlamesa.medium.com/atlantas-virtual-academy-is-a-child-rights-catastrophe-df2a32afa0ad | Social Policy, Education, Politics Reporter | 15400 | 5 | 1 |
| 7/25/22 | If you doubt that the #monkeypox outbreak is driven overwhelmingly by sex between men or that there is somehow a shadow epidemic among women and kids and that it will run rampant in schools and public bathrooms, I urge you to check out all the mounting evidence to the contrary. | Health and Science Reporter | 17700 | 34 | 6 |
| 7/21/22 | For everyone who is worried that #monkeypox is going to tear through society, upending elementary schools left and right, this finding from an important new @NEJM should come as reassuring news. That is, unless some people out there *want* to live in constant fear and hysteria... | Health and Science Reporter | 17700 | 13 | 0 |
| 7/20/22 | I don't think school districts should be worrying about monkeypox. A problem during the pandemic was that school districts took it upon themselves to become little public health departments -- APS even hired an epidemiologist! Let public health workers worry about public health. | Social Policy, Education, Politics Reporter | 15400 | 12 | 1 |
| 7/18/22 | Unless the reopening of schools & colleges leads to a surge of sex between males, I can't see how that change would have any effect on the #monkeypox outbreak. Experts agree that the outbreak is driven by sex between men. Monkeypox does not appear to transmit readily via the air. | Health and Science Reporter | 17700 | 11 | 0 |
| **PhD, MPH, other Ed. degree** |  |  |  |  |  |
| **Date** | **Tweet** | **Author Credentials** | **Follower Count** | **Likes** | **Retweets** |
| 9/10/22 | Monkeypox cases are going down in USA and EU. Test positivity rates declining. No signs of spread in schools or day care centers. https://ourworldindata.org/monkeypox | PhD Professor Virologist | 23700 | 1,135 | 228 |
| 8/21/22 | Very irresponsible reporting. There is no evidence that #monkeypox will spread in schools. Actually, the evidence argues exactly the opposite is true. | PhD Professor Virologist | 23700 | 768 | 124 |
| 8/21/22 | Because fear mongering about schools for clicks has been the MO for the @nytimes for the last two years. If they can't do it for Covid anymore, monkeypox is the next best thing. | PhD Infectious Disease Scientist | 16300 | 83 | 10 |
| 8/4/22 | This is why it’s frustrating AF to see people on here talking about how school isn’t gonna be safe with the monkeypox outbreak. It’s just fear-mongering for the sake of fear-mongering. | PhD Ethnomusicology | 8894 | 6 | 0 |
| **JD** |  |  |  |  |  |
| **Date** | **Tweet** | **Author Credentials** | **Follower Count** | **Likes** | **Retweets** |
| 8/9/22 | And so it begins: 'Colleges, univ's in Illinois begin monkeypox prep.'  #PermanentEmergency #Illinois #schools #Tuesday  @vjeannek https://chicagotribune.com/news/ct-monkeypox-universities-colleges-20220809-yb3qayt3kbhdpg25cik67gz63e-story.html | JD | 9481 | 12 | 0 |
| 8/6/22 | Close schools for 18 months out of an abundance of caution, but just practice slightly safer sex to avoid monkeypox. WTAF. We have destroyed children for 1/1000 of the risk that monkeypox poses to adults. Public health is so broken. | JD | 1647 | 19 | 2 |
| 8/4/22 | There have been, to date, zero monkeypox deaths in the US. Yet there are people calling for school shutdowns, monkeypox vaccine requirements, and all sorts of other so-called disease avoidance measures. This is what happens when a society elevates virus avoidance above all | JD | 114600 | 2,132 | 443 |
| 5/22/22 | In 2003, 47 confirmed cases of monkeypox were reported from six state...All people infected with monkeypox in this outbreak became ill after having contact with pet prairie dogs  Mask up, pivot to iPad school & mail in ballots!  Monkeypox in the US | JD | 25300 | 32 | 5 |

**Table S3.** All tweets categorized as inaccurate/exaggerated. Bolded text in the Tweet column is added notes by the investigators. Tweets that are comments are denoted with a bold (comment). Tweets that were deleted are denoted with a bold (deleted tweet).

| **Health Care** |  |  |  |  |  |
| --- | --- | --- | --- | --- | --- |
| **Date** | **Tweet** | **Author Credentials** | **Follower Count** | **Likes** | **Retweets** |
| 9/15/22 | We have the first case of #MonkeyPox in orange county school, FL. Activities involving prolonged Skin-to-Skin contact with an infected individual, such as wrestling, tackle football, prolonged hugging, and kissing, are the risk factors for Monkeypox in K-12 students. | MD | 8826 | 13 | 9 |
| 9/12/22 | Schools are petri dishes. Covid, monkey pox, now polio threat. The best parents cano is make sure kids immunizations are up to date and get kids to wear a mask | Retired RN | 6309 | 1 | 0 |
| 9/10/22 | Not surprising to hear monkey pox spreading around schools. So sad | RN | 316 | 1 | 0 |
| 9/9/22 | Was glad to get my #monkeypox vaccine at  @NCCU yesterday. As we start a new school year, it’s important for all of us to be sure we’re up to date on our vaccines. | NC Department of Health and Human Services Secretary | 4980 | 15 | 3 |
| 9/5/22 | $TOMDF - pharmacies getting ready COVID+MonkeyPox dual screening coming for school outbreaks in months ahead. COVID At-Home COVID PCR testing will also be huge. Marketing push coming for consumers of antigen (COVID PCR saliva 10/25/50 packs 36h TAT) & schools (MPX +COVID saliva). | CEO Todos Medical and Executive Chair at Amarantus Bioscience | 3995 | 31 | 8 |
| 9/4/22 | Monkeypox cases plateauing/trending down overall, but the proportion of cases unable to be traced to men who have sex with men is rapidly increasing. Also schools and sports starting back up nationwide. The next few weeks will be telling. This could fizzle out, or explode… | Former Surgeon General MD MPH | 80300 | 161 | 29 |
| 8/30/22 | School in Texas has just started and there are already two monkeypox cases. Watch it explode under the anti-science #Republican lack of leadership, because they don't care who gets sick or who dies. | Retired RN | 2573 | 1 | 0 |
| 8/26/22 | My Friday gift is: the 5TH ANNUAL #SHOTWAVE! It's simple: #GETVACCINATED: ANY vaccine (school, flu, monkeypox, COVID). Post a pic of your bandaid to me here or (new!) Instagram. I donate  @UNICEF  so kids worldwide can #getvaccinated & we create a #SHOTWAVE. https://gofundme.com/f/shotwave-2022 | MD PhD MPH | 9911 | 13 | 9 |
| 8/24/22 | What were they saying again about monkeypox not spreading in schools? | MD | 4222 | 14 | 3 |
| 8/23/22 | College Kids and Pet Owners Should Beware #Monkeypox Too  The outbreak has proved more persistent than many thought, and governments have been slow to roll out vaccines. We can’t lose vigilance in the back-to-school rush. https://bloomberg.com/opinion/articles/2022-08-23/monkeypox-outbreak-college-kids-and-pet-owners-should-beware-the-virus-too | MD Professor | 4804 | 1 | 1 |
| 8/21/22 | HIV was a disease of men having sex with men until it wasn’t. In Africa monkeypox spreads in schools. We don’t know if that will happen in the West, but we also don’t know that it won’t #HygieneMatters | MD PhD | 11000 | 10 | 6 |
| 8/20/23 | 🇺🇸 has 1/3 of #Mpox cases & ? #vax available. #School starts! | RN | 6,549 | 1 | 0 |
| 8/18/22 | Children with monkeypox: This is the tip of the iceberg (as symptoms can be mistaken for other rash in kids) & we expect the numbers to rise. With school opening & shortage of vaccines, these numbers will ⬆️in the fall unless we expand testing & vaccine. | MD MPH Professor | 47100 | 0 | 5 |
| 8/17/22 | ASD schools start tomorrow. How are we preparing knowing there are no guidelines or strategies in schools or the community to keep kids, staff, and teachers safe from simultaneous surges in SARS2, Flu and Monkeypox? | PhD MPH CPH RN | 1038 | 14 | 3 |
| 8/17/22 | #Monkeypox has likely descended upon UMD —and this campus almost certainly won’t be the only one. Schools are not prepared.   The to-be-overhauled CDC needs to jump on this problem STAT and issue clear, actionable guidance. | MD Professor | 8453 | 2 | 0 |
| 8/16/22 | We have a 2 yo and a dog positive for Monkeypox now. I'm concerned this may spread like crazy in schools and daycares. | MD PhD | 2766 | 4 | 0 |
| 8/16/22 | #Monkeypox has already spilled out of the #LGBTQ community. We have reported cases in kids, women, & now dorms. With schools opening soon, we have to find ways to vaccinate groups of people that cluster together for prolonged periods of time.  @VictorBlackwell @AlisynCamerota | MD MPH | 5990 | 11 | 6 |
| 8/16/22 | Thank you, My Friend🙏🏻🙏🏻🙏🏻  So, back to normal programming for this Pandemic Foot Soldier🦠🦠🦠  MPox spreading in schools in the USA.   Clearly, this is going to be the next major issue to confront Australia 👇🏻👇🏻👇🏻👇🏻👇🏻 | MD PhD | 9,728 | 11 | 4 |
| 8/16/22 | Taps Mic:  @CDCgov. In addition to reinstating basic guidelines for mitigation practices within schools for COVID, now would be a good time to change messaging on #Monkeypox stating “a virus does not care about age, gender, nor sexual orientation”. | MD | 62300 | 123 | 48 |
| 8/14/22 | I comment on @FOX5Vegas the recent reports of a #monkeypox case in one of our local high schools  It was only a matter of time before cases started to pop up in "high congregate" settings like schools  Vaccine Supply is limited but available 4 at risk  #TwitteRx #MedTwitter | Pharmacist | 836 | 2 | 2 |
| 8/14/22 | Schools should have clear guidance for staff and students on monkeypox that distinguishes likely and unlikely risks, encourages vaccination appropriately for those at more risk, avoids unnecessary euphemism and tackles fears | Infectious Disease (anonymous) | 45200 | 8 | 2 |
| 8/12/22 | I literally only saw 2 other people wearing masks in the school I work in. There’s both Covid & monkeypox to worry about now and apparently this country’s just giving up on any kind of prevention/mitigation measures? This is just bad policy that’s going to cause long-term harm | Psych Doctor | 589 | 3 | 0 |
| 8/12/22 | One way MPX [monkeypox] spreads is through skin to skin contact. School is starting. Anyone ever see impetigo outbreaks among athletic teams? Wonder what could happen this school year if we ignore this one b/c " we don't have gay kids on our football team" [[sarcasm alert]] | NP | 5612 | 40 | 4 |
| 8/11/22 | The #CDC has decided that since America has moved past the #pandemic, they might as well give the public what they want. All of this w/ schools reopening & #monkeypox cases climbing! | MD MPH | 5990 | 24 | 1 |
| 8/11/22 | 30 days ago, there were 767 documented monkeypox cases in the U.S. We now have crossed the 10,000 mark - those r just the cases we know about. Without more testing & more vaccines, I am skeptical of our ability to contain this.The real question is what happens as schools reopen. | MD JD | 8011 | 85 | 21 |
| 8/11/22 | genuine question. what are schools doing to prepare for monkeypox this fall, esp K-12?  chatting with teacher friends who are feeling both anxious and unprepared for monkeypox in the classroom. | MD Student | 7293 | 25 | 1 |
| 8/11/22 | Given that children are at highest risk of complications due to monkeypox and also least able to tolerate ACAM2000 and are forced in close contact at daycare/school what is the plan to protect them? Will we be saving Jynneos doses for when this spreads to schools?  @PTF_org **(comment)** | MD | 1955 | 75 | 12 |
| 8/10/22 | Covid. Monkeypox. Flu. ??? School starts over the next the couple of weeks.  Please send tots and pears to your pediatrician friends 😭😭😭 #pandemic #Tweetiatricians | DO | 917 | 7 | 0 |
| 8/10/22 | Small, college towns will soon understand the protective bubble they live in once schools restart, and students return.  Surveillance testing, contact tracing, and post-exposure vaccination for #Monkeypox will be key given the weeks long incubation period. | MD | 62300 | 36 | 9 |
| 8/9/22 | Everyone is rolling back restrictions including the #CDC as #kids go back to school. Add #monkeypox to the threat now.  But you don’t have to. Back to the basics - hand washing & #masks | MD MPH | 5990 | 18 | 4 |
| 8/8/22 | When you pen an op ed saying #monkeypox cases are doubling every two weeks… and a few hours later you find out they are now tripling…  🙊 🚀   The monkeypox emergency is going to affect schools, colleges. Be ready | Former Surgeon General MD MPH | 80300 | 374 | 113 |
| 8/8/22 | “There is definitely potential for spread of monkeypox” in daycares, schools, college campuses, prisons, and other similar settings, said Dr. Alexandra Brugler Yonts, an infectious disease specialist at Children’s National Hospital in Washington, D.C. | MD | 59700 | 183 | 84 |
| 8/7/22 | Monkeypox at a daycare was 'only a matter of time,' expert says. Next up: pools, sports, schools - FORTUNE **(quoting Fortune article)** | MD | 30400 | 1 | 1 |
| 8/7/22 | VAX ALREADY APPROVED FOR AFFECTED CHILDREN! Monkeypox at a daycare was ‘only a matter of time,’ expert says. Next up: pools, sports, schools #NewsBreak **(quoting Fortune article) (deleted tweet)** | RN | 2818 | 1 | 0 |
| 8/7/22 | Monkeypox at a daycare was 'only a matter of time,' expert says. Next up: pools, sports, schools \| Fortune  “Anywhere that close physical, skin-to-skin contact occurs—particularly of people who are in various stages of undress—there is risk,” **(quoting Fortune article)** | Surgeon | 937 | 1 | 0 |
| 8/6/22 | I am worried about monkey pox when school reopens. Curiously, I haven’t heard any plans from state or local school district to anticipate and prevent it 😭 | MD PhD | 6057 | 1 | 0 |
| 8/5/22 | I don’t want to assume, but I’m guessing all the kids at the day care center don’t identify as gay men.   Monkeypox initially was spreading in the MSM community, but it is NOT a “gay” disease. Expect to see more of this (school/ daycare/ prison cases) in the coming weeks. | Former Surgeon General MD MPH | 80300 | 716 | 163 |
| 8/5/22 | As predicted, Monkeypox started as an STI, but it’s not staying that way. What does this mean? Time to plan to prevent not one, but two dangerous viruses from spreading in schools and daycares. Denying this unfortunate reality will not make it go away. | MD | 38400 | 460 | 159 |
| 8/5/22 | All of #MedTwitter has worried about this, but I hoped it would not happen so soon. I hope every childcare facility & school system is thinking now about how they will handle a #monkeypox exposure or outbreak. | MD | 60300 | 211 | 69 |
| 8/4/22 | So NOW MonkeyPox is a health emergency, huh.  Turns out everyone closing their eyes and screaming “It’s a GAY disease” doesn’t actually reduce transmission.  Schools start back in a week and there’s a highly transmissible disease AGAIN. With no vax available  Fucking ridiculous. | MD (anonymous) | 833 | 1 | 0 |
| 8/4/22 | And toss in Monkey pox for extra fun because if it hits school I'm done … | RN | 386 | 1 | 0 |
| 8/4/22 | The @CDCgov wants to lift Covid restrictions in schools just as Monkeypox starts spreading in schools. We are the most resourced idiotic country when it comes to managing the public health. Happy Back to School! | MD | 15900 | 16 | 2 |
| 8/3/22 | So hey, #monkeypox. Gonna be like chickenpox, but EVERYONE in the family gets it at the same time? Like, all the school kids and their parents and sibs? And 2-4 week isolation for everyone? Am I missing anything here? #WelcomeBackToSchool | DO | 3629 | 1 | 0 |
| 8/3/22 | There's so many what ifs surrounding #MonkeyPox, including women, children returning to schools in few weeks from PreK to University. | Retired RN | 3000 | 1 | 0 |
| 8/3/22 | Is anyone else concerned about monkeypox and covid transmission at Outside Lands in SF, Sturgis in SD and Burning Man in Nevada this month? And other gatherings. And school getting back in session. Just me? Okay 😵‍💫😳😷 | MD | 2430 | 60 | 13 |
| 8/3/22 | As we inch closer & closer to school reopening, I get more concerned about a lack of a standard response plan...This is my county... More Monkeypox Cases Detected In Camden County | NCSN | 12300 | 8 | 0 |
| 8/2/22 | I hope the schools are ready for Monkey Pox because this isn't a drill it's here !!! | RN | 386 | 1 | 1 |
| 8/2/22 | Radical Tim Michels (running for governor in WI) has ads saying if elected he will “keep schools open 5 days a week no matter what Fauci says!” Running on a promise to ignore health experts is so on brand for Republicans. COVID, monkeypox, polio, be damned. | RN Professor | 669 | 16 | 5 |
| 8/2/22 | #Schools should STAY OPEN this fall. Schools should also bring back the mask mandates, improve air circulation and embrace outdoor learning. I would also love #testtraceisolate to come back, both for COVID & for Monkeypox. We can stay open with precautions in place. | Pharmacist | 9266 | 216 | 33 |
| 8/2/22 | Monkey pox is gonna go crazy once the kids go back to school | RN | 1278 | 4 | 0 |
| 8/2/22 | Going into a school year with the pandemic and monkey pox on the rise feels pretty terrifying.   I’ve seen how we public health… | Psychotherapist | 11300 | 138 | 16 |
| 8/1/22 | Looking forward to the first few weeks after school starts for the physicals/forms to slow down.  Absolutely not looking forward to the rest of the year after that. Brace yourselves my #tweetiatrician friends.   #COVID19 #MonkeyPox #ColdandFluSeason | MD | 15900 | 8 | 3 |
| 8/1/22 | #monkeypox cases in US up to 5800. This there is community spread now. School are starting we need to have plan for schools and universities. Need to screen students for rash and infection symptoms. Need more public health messaging. Need vaccines readily available. | MD Professor | 6636 | 22 | 4 |
| 8/1/22 | How are schools, colleges, daycares and workplaces going to stay open with uncontrolled monkeypox and Covid spread? Anyone have a plan? **(deleted tweet)** | MD | 4363 | 148 | 23 |
| 7/31/22 | Maybe school start should be delayed until we have our shit together ? What about til after labor day ? Ya know in interim make vaccines for kids very easy to get and come up w plan for monkeypox including massive vaccine production and distribution campaign …? It makes sense | MD | 3708 | 31 | 3 |
| 7/31/22 | What keeps me up at night is thinking about how monkeypox PLUS Covid is going to impact kids this fall/winter.  Covid has already turned daycare into a hot mess and resulted in children missing 1.5+ years of in-person school. | MD Professor | 3895 | 50 | 17 |
| 7/31/22 | Monkeypox is Spreading Fast. Now Kids Are Getting It, Too - Bloomberg. “Already, more than 80 kids across several countries have contracted monkeypox” Perhaps before schools open this Fall we should be offering vaccines before it is too late? | MD PhD | 18700 | 73 | 26 |
| 7/31/22 | Time to ask @CPHO @CDNMinHealth @JustinTrudeau what their plan is to contain #monkeypox in Canada. Vaccines? Testing? In areas where #monkeypox is endemic, the majority of cases are in those under 15. What’s the plan to protect us and our children before schools open?  #Canada | MD | 15400 | 70 | 25 |
| 7/31/22 | Monkeypox is going to enter your children's daycares and schools. Covid did and you did not protect your children. It will not stop with mpx. It will get worse. You are responsible for what happens to your children. | Retired RN | 945 | 5 | 0 |
| 7/30/22 | Get all the kids vaccinated for monkeypox. We need to keep the schools open, and this will be a highly vulnerable group re transmission. | Physician | 17400 | 12 | 2 |
| 7/30/22 | In the current, Trump-inspired environment of public mistrust in science & the medical community, monkeypox wasn't taken seriously at the start, when it could've been fully contained. Now, just wait until it enters the pediatric population. Daycares, schools will have to close 🙄 | Physician | 17400 | 11 | 4 |
| 7/27/22 | THIS MATTERS!  Monkey pox is currently spreading via close sexual contact in populations of men who have sex with men  IT IS ALSO SPREAD by any close contact, droplets, and fomites  The way we’re discussing it as an STD is going to get gay people killed when kids get it at school | Medical Student | 6001 | 2 | 0 |
| 7/28/22 | Schools were to be kept open SAFELY. Please provide update on how you will accomplish this, given the more contagious BA.5 and now Monkeypox. Your silence implies you have NO PLAN to keep kids safe. @POTUS @PTF_org @AAPPres @ASlavitt @ashishkjha @WHCOVIDResponse | DO | 488 | 103 | 34 |
| 7/26/22 | People want to downplay this while sitting on stores of the small pox vaccine. Just make it available and let folks get it before school starts… | MD (anonymous) | 4231 | 1 | 0 |
| 7/26/22 | I just thought about what would happen if a student came to school with monkeypox.  Y'all, this is going to get bad when schools are open. | Speech Language Pathologist | 2024 | 26 | 0 |
| 7/25/22 | With the kids going back to school in a little over a month, I’m terrified of monkey pox. Vaccination is abysmal and they’re treating it like an STD. We’re so screwed. | PT | 3411 | 4 | 1 |
| 7/25/22 | Please correct anyone and everyone who says monkeypox is a sexually transmitted infection.   It matters because it could very well end up in a school setting and the hate & lies about how they got it will be adding trauma to yet another unchecked infection. | PT | 14000 | 16 | 3 |
| 7/25/22 | I would call them "closures" but anyone who thinks schools will be open for all of this fall/winter is really not understanding monkeypox and Covid and what they will do to schools. **(deleted tweet)** | MD | 4361 | 16 | 5 |
| 7/24/22 | Wastewater testing for Monkeypox needs to happen ASAP in all jurisdictions. In these uncertain times, you need as much information as possible, especially with the school year starting soon. | MD | 38400 | 561 | 180 |
| 7/23/22 | Not ready for Monkeypox season to hit just as my kids go back to school | MD | 6330 | 11 | 0 |
| 7/23/22 | Schools are starting in a couple of weeks and we have rising COVID and now #monkeypox What are we doing to mitigate?? | MD Professor | 2945 | 5 | 1 |
| 7/23/22 | Shit is about to go more sideways than anyone realizes with this. Think about chicken pox outbreaks in primary schools and daycares. Now remember the adults also don't have protection with narrow exceptions. We have entered the witch hunt phase **(tweet is referring to Mpox, quotes a tweet about Mpox)** | PharmD | 537 | 1 | 1 |
| 7/23/22 | About time 😬 and with now further confirmed cases in Children, is #Belgium preparing for the upcoming opening of our schools? #monkeypox #PHEIC  @BenWeyts   ➡️"respiratory secretions"=droplets/aerosols! #airbornepossibilities #ventilation  https://cdc.gov/poxvirus/monkeypox/transmission.html | Surgeon | 4998 | 13 | 2 |
| 7/23/22 | We have a short opportunity to try to catch up to monkeypox. @potus once he’s better from Covid and  @cdc could activate multiple large factories to help produce sufficient ppe/vaccine 💉 to prepare the front line/essential workers. Then school children. Then adults. | MD | 1207 | 2 | 0 |
| 7/22/22 | MONKEYPOX IS NOT A SEXUALLY TRANSMITTED DISEASE!  It is a contact-transmitted disease.  Since it's first outbreak was among gay men, & sex is a contact-rich behavior, it currently *masquerades* as an STD. But *even non-sexual contact* can spread it.  GRADE SCHOOLS ARE VULNERABLE. | Retired MD (anonymous) | 3129 | 31 | 18 |
| 7/22/22 | Children play with children 🚨 #CDC needs an urgent #MonkeyPox containment plan for the next school year👇🏼👇🏿👇🏾👇🏽 | MD | 660 | 1 | 0 |
| 7/22/22 | Monkeypox, if not rapidly contained, will inevitably spread among kids in daycares, schools, and sports leagues. The vaccine currently being used won’t be the silver bullet since supply is limited and it wasn’t studied in children so it’s not approved for <18s. What’s the plan? | MD Professor | 65300 | 3,683 | 1,332 |
| 7/22/22 | Monkeypox will become established in the pediatric and general population and will transmit through daycares and schools. It is not an STD. It is like MRSA. This isn’t rocket science. **(deleted tweet)** | MD | 59700 | na | na |
| 7/21/22 | Looking at these monkeypox numbers in NYC and dreading what happens once schools open up. | MD MPH | 23900 | 3 | 2 |
| 7/21/22 | As mentioned already; when MonkeyPox gets into the schools, it will spread like wildfire—- How soon after until  @POTUS  is blamed?  @CDCDirector @CDCgov @jeremyfaust @CarlosdelRio7 | MD MPH Professor | 3378 | 2 | 2 |
| 7/20/22 | Heads up!!! Mask up! Get your #influenza vax as soon as it comes out. Get your booster(s) for #covid now. Learn how #monkeypox transmits. There is the potential for ##BA5, #BA275, flu and monkeypox to all surge/peek about the time kids go back to school. What could go wrong?🤪 | PhD MPH CPH RN | 1038 | 1 | 2 |
| 7/20/22 | Things that'll probably be a bad idea (and/or have their economies crater) once monkeypox hits, a casual list:  Gyms Restaurants (even to go) Malls/shopping in person Schools  Grocery stores Hotels (all travel, really)  I'll add more as they come to me… | RN | 5264 | 82 | 22 |
| 7/20/22 | Kids going back to school is going to make Monkey Pox everyone’s problem **(deleted tweet)** | MD | 6300 | na | na |
| 7/20/22 | Just waiting for monkeypox to show up in a high school wrestling tournament. | MD | 59700 | 1,240 | 139 |
| 7/17/22 | The big question: what will happen when school children in the US are exposed to monkeypox? The answer is that right now, we don’t know https://med.stanford.edu/news/all-news/2022/06/monkeypox.html | MD PhD | 11000 | 4 | 1 |
| 7/14/22 | I hate to tell you all this, but #covid19 is still a pandemic, and now #monkeypox is too. And both are gonna get a LOT worse before they get better… just wait till schools - including colleges- reopen in a few weeks… 🤦🏽‍♂️ | Former Surgeon General MD MPH | 80300 | 661 | 343 |
| 7/7/22 | Things I'm not looking forward to:  School nurses calling me to report #monkeypox outbreaks on their school wrestling teams  Cause 10/10 this is the direction we are heading | Nurse (anonymous) | 7252 | 120 | 19 |
| 6/28/22 | What's the plan in your country to avoid monkeypox in schools?  Children are a vulnerable group.  According to  @who  guidelines:  Need for masks in close contact, transmission by shared surfaces/objects possible  This must not enter schools. | MD MBA MPH | 21100 | 202 | 104 |
| 6/7/22 | Just wait until monkeypox hits the schools and daycares. The vast majority of fatalities in monkeypox occur in children. | MD | 59700 | 838 | 237 |
| 6/3/22 | You heard it here first. A journey into 12 months from now. America: "There was no way we could have predicted monkey pox would spread. It's now endemic and we all need to return to normal for the good of the economy. We need to open schools up." | RN | 2612 | 9 | 0 |
| 5/27/22 | All kids should have their rashes checked out. All kids.   This disease is serious for kids. Any kid could be the index case in childcare and school.   #ProtectKids #monkeypox **(comment)** | MBBS BSc | 35800 | 32 | 3 |
| 5/24/22 | Why should we bother with preventing #COVID19 transmission or #monkeypox when we just allow 14 children die while in school? It’s gross #nopriorities | Physician | 464 | 3 | 0 |
| 5/20/22 | So what's the plan  If monkeypox enters schools? Kindergartens?  'oh it's just chickenpox' won't work because it isn't chickenpox | MD MBA MPH | 21100 | 157 | 56 |
| 5/19/22 | So how many monkeypox outbreaks are going to happen in schools before we close them until we can vaccinate the kids? | MD | 59700 | 1,584 | 219 |
| 5/19/22 | an unsolved issue is what to do if monkeypox contacts are children. there's no approved vaccine under age18.   how is your country approaching this problem? | MD MBA MPH | 21100 | 375 | 92 |
| **Health Reporter** | |  |  |  |  |
| **Date** | **Tweet** | **Author Credentials** | **Follower Count** | **Likes** | **Retweets** |
| 9/5/22 | Half of the interstate system in the eastern US right now is clogged up with congestion from the holiday weekend. In the northeast schools if it has not already started starts for the remaining kiddies. Good luck in there not being a massive covid/monkeypox surge next few weeks. | COVID Reporter | 16500 | 29 | 3 |
| 8/29/22 | Georges C. Benjamin, exec director of  @PublicHealth , said it's inevitable monkeypox will surface on college campuses this fall: "... all schools should assume that they’re going to have somebody on their campus that has monkeypox." Is Florida prepared? | Health Reporter | 498 | 7 | 4 |
| 8/24/22 | Everyone is talking about student loan debt relief instead of elementary school students with monkeypox and kids dying of Covid. | Medicine/ Law | 50300 | 586 | 99 |
| 8/15/22 | 😳 Why aren’t pediatric #monkeypox cases being widely reported??   @CDCgov  - how do parents protect their kids when school starts? | Health Reporter | 30200 | 265 | 82 |
| 8/15/22 | You know people are completely delusional about the current situation, because when you ask them how they plan to keep their kids safe during the upcoming school year, they say, “Oh, we already had Covid, & our family isn’t at risk for monkeypox.” | Medicine/ Law | 50300 | 1,376 | 267 |
| 8/13/22 | Parents are sending kids back to school, believing with their whole hearts kids can’t get monkeypox & they won’t get Covid anymore either. The Biden admin is focused on midterms, but mass infecting kids with Covid AND monkeypox isn’t a winning strategy. Your advisors are lying. **(deleted tweet)** | Medicine/ Law | 50300 | 1,559 | 326 |
| 8/12/22 | NEW: Clark County — that's Las Vegas, for non-Nevada folks — announces a monkeypox case at a local high school.  Unclear whether it's a teacher, student, staff, etc. but underscores some of the concerns about this spreading in a college or high school setting. | Health Policy Reporter | 30000 | 20 | 9 |
| 8/10/22 | If you are able to keep your child home from school or daycare, this is certainly the safest option.  We must & will advocate for layers of protections in schools & all indoor spaces.  But if you *can* keep your kids home while COVID-19 & monkeypox are circulating, I support you. | Evidenced Based Policy | 3175 | 8 | 4 |
| 8/7/22 | Also what will the public say when they realize that every human being can get monkeypox on top of that. I feel so bad for what the kids are about to go through with schools opening again. **(comment)** | COVID Reporter | 16500 | 42 | 1 |
| 8/6/22 | Because children have not been the primary demographic affected outside endemic countries, we havent talked nearly enough about monkeypox and children, esp w/ school coming up. Historically, children have had the most severe outcomes from the virus | Health Science Writer | 15000 | 9 | 2 |
| 8/4/22 | “No one knows full extent of outbreak after delays with testing, data gathering and vaccine rollout allowed monkeypox to spread in US”   As we reopen schools, we have the world’s largest monkeypox outbreak. | Medicine/ Law | 50300 | 220 | 95 |
| 8/4/22 | Health experts: Fall school semester could amplify spread of monkeypox **(quoting local article)** | COVID Reporter | 16500 | 22 | 8 |
| 8/1/22 | Monkeypox isn’t the only virus posing a threat to children as the school year rapidly approaches.   Covid has been quietly doing push-ups, bulking up, (while still mutating!) & preparing to disable & kill even more of the nation’s children. | Medicine/ Law | 50300 | 868 | 319 |
| 7/29/22 | #Monkeypox is Spreading Fast. Now Kids Are Getting It, Too. The virus could become hard to contain in schools, #childcare settings, by  @g0ingmad  https://bloomberg.com/news/articles/2022-07-29/as-monkeypox-spreads-kids-can-get-monkeypox-too#xj4y7vzkg… via  @business   @PublicHealth  #pediatrics #MPXV | Health Columnist | 137200 | 32 | 18 |
| 7/27/22 | Children under eight are considered among those at higher risk for serious infection for monkeypox. Emory’s  @CarlosdelRio7  told  @11Alive : “It’s very possible that cases in children are going to increase.” | Emory University | 5404 | 11 | 9 |
| 7/26/22 | So, monkeypox—a virus that can cause blindness, disfigurement, extreme complications, & death—has been reported on at least 5 college campuses & in county jails. It’s also infecting kids. There is no plan for fall/winter. We cannot accept doing nothing. | Medicine/ Law | 50300 | 126,000 | 33,502 |
| 7/25/22 | Teachers and kids will be out with monkeypox and covid schools will close because so many staff are sick with one of our many viruses. **(comment)** | COVID Reporter | 16500 | 3 | 0 |
| 7/18/22 | So currently there are no public health measures in the local schools. Covid is ignored. Monkeypox is ignored. No one masks. I’m expected to just suck it up, send my kids, & hope for the best. There are NO accommodations for high-risk families. This is sick. It’s not normal. | Medicine/ Law | 50300 | 3,755 | 869 |
| **PhD, MPH, other Ed. degree** |  |  |  |  |  |
| **Date** | **Tweet** | **Author Credentials** | **Follower Count** | **Likes** | **Retweets** |
| 9/19/22 | When an #Airfilter is so cheap to run, why wouldn’t we protect schools @kitmalthouse?  #Asbestos #Covid #Monkeypox etc. #UK does not have to be world leader of 1) illness through school and 2) absence through avoidance | PhD Professor Environmental Scientist | 3190 | 2 | 2 |
| 9/9/22 | Finally some promising news about monkeypox. I just hope that with schools starting this doesn't take off again. | Epidemiolgist | 15200 | 57 | 6 |
| 9/8/22 | First monkeypox case in an Arkansas school. Going to be a semester of "fun" scrutinizing symptoms, with monkeypox risk & the much-more-common COVID going around. | Epidemiolgist | 2759 | 19 | 12 |
| 9/7/22 | As school just starts to ramps up, so are monkeypox cases in kids.  Thread 👇 | Scientist | 1316 | 7 | 3 |
| 9/2/22 | Great thread on another awful school year ahead. This time with polio and monkeypox on the side. **(deleted tweet)** | PhD Professor | 61200 | 58 | 24 |
| 8/31/22 | NEW—3 children have tested positive for #monkeypox in Dallas County. Just ignore this you think it’s a total coincidence that schools have started to reopen. | Epidemiolgist, ScD | 783100 | 1,027 | 455 |
| 8/29/22 | "There have been no documented cases of monkeypox transmission in schools during this outbreak" isn't as reassuring as you think it is.  Most cases of monkeypox right now are in Europe and the USA.  Schools there have been out for summer during much of the outbreak. | PhD Professor Evolutionary Biology | 80200 | 342 | 41 |
| 8/28/22 | I keep hearing on  @citynewscalgary  (660) that nobody needs to worry about #monkeypox spreading in schools because it's mainly transferred by skin contact... anyone who thinks skin contact doesn't happen between school age kids seriously doesn't have kids. | PhD BSc | 1161 | 4 | 1 |
| 8/28/22 | @CanadianPress -Jason Kindrachuk, @umanitoba virologist “urging Canadian universities and colleges to be proactive about preventing #monkeypox from spreading on campus…says schools should be raising awareness about the risks …as students prepare to come together this fall” | PhD Student | 4261 | 1 | 1 |
| 8/21/22 | What are High School/University Wrestling programs doing to prevent the spread of MonkeyPox this Fall??? | Parasitologist | 6411 | 19 | 2 |
| 8/21/22 | Who will wait to see if someone else will take care of this problem, and who will act?  Monkeypox may be on its way to a school near you **(quoting Hill article)** | PhD Physics | 93100 | 290 | 126 |
| 8/30/22 | Back-to-school is upon us, amidst a monkeypox pandemic, rolling Fascist coup, insurgency, and GQP war on LGBTQ ppl. CDC still treating mpx as a 'gay STI,' but quietly admits virus is spread on surfaces and is also airborne. Volatile confluence. 🧵 1/ | PhD | 4673 | 3 | 3 |
| 8/19/22 | #Monkeypox may be on its way to a school near you **(quoting Hill article)** | PhD Psychology | 1360 | 20 | 31 |
| 8/17/22 | I'm a graduate of Columbus Public Schools and support these teachers. Teachers, students and staff deserve buildings and classrooms conducive to learning and with good ventilation especially in these times of Covid (and Mpox) | PhD Professor | 3,514 | 6 | 2 |
| 8/16/22 | 5yo starts K soon, which should be exciting milestone. Instead, constant dread given CDC guidance downplaying masking/testing, antiquated school ventilation system, lack of access to new Covid booster for kids, monkeypox, & risk to infant sibling. Choice is education OR health. | PhD Ecologist | 2248 | 11 | 0 |
| 8/16/22 | STOP FAULTING MSM—The first step to stopping #monkeypox is admitting / recognizing that transmission can be non-sexual. There is risk even at **crowded outdoor** events. Once you recognize this—we need to wake up about imminent #MPXV school transmission & need for mitigation. | Epidemiolgist, ScD | 783100 | 522 | 259 |
| 8/15/22 | Monkeypox case confirmed at a Las Vegas-area high school. Unclear if student, teacher or staff. 75 other monkeypox cases in the region. Doctors expect more #monkeypox to emerge as schools reopen. | Epidemiolgist, ScD | 783100 | 1,137 | 609 |
| 8/14/22 | My advice for parents this school year.   Wait for 2 weeks after the 1st day of school. See how bad Monkey Pox and Covid is destroying all the kids.   Reassess at week 2… | Doctor (anonymous) | 10400 | 605 | 141 |
| 8/12/22 | * Opens email *  Kids’ school:  “Monkeypox information” * Closes email *  Sigh. | PhD MPH | 4867 | 9 | 0 |
| 8/11/22 | Impeccable timing,  @CDCgov ! Framing “normalcy” as “no longer needing quarantine/ testing restrictions, will make it harder to control monkeypox when it spreads in schools. And the “if you don’t test it doesn’t exist” strategy won’t help when the kids all show up with MPX pustules | Biologist | 5629 | 11 | 3 |
| 8/11/22 | Disinfo like this from Reuters obscures the fact that monkeypox includes airborne/respiratory transmission. With CDC and Fascist GQP promoting the lie that mpox is a "gay STD," MSM is setting the stage for MAGAt violence as back-to-school kids become airborne-infected w/ mpox. | PhD | 4673 | 4 | 2 |
| 8/10/22 | We need vaccine-plus strategies NOW. Especially as monkeypox is spreading.  @jjhorgan if you care about economic & physical well-being of BC: immediately adopt masking strategies, ventilation overhauls in schools+public spaces, adequate paid sick days + provide adequate housing **(deleted tweet)** | PhD | 35700 | 14 | 0 |
| 8/10/22 | Monkeypox at a day care was ‘only a matter of time,’ expert says. Next up: pools, sports, schools **(quoting Fortune article)** | MPH | 303 | 2 | 1 |
| 8/8/22 | Im so excited to start tracking monkeypox at work. Like Im geeked to be running point on this. But also highly annoyed at my own shithole state refusing to allocate vaccines to the general public (teachers, healthcare providers, etc.). Mind you, school starts this week. | MPH | 17700 | 23 | 3 |
| 8/8/22 | I’m going to add to this that the isolation time for MonkeyPox is 3-4 WEEKS. If you can’t afford to take 3-4 weeks off of work, you should send your kids to school in masks. | M.Ed | 387 | 6 | 0 |
| 8/8/22 | #Monkeypox at a daycare was ‘only a matter of time,’ expert says. Next up: pools, sports, schools: https://tinyurl.com/5bkujsd2 #publichealth #MPV #MPX #education **(quoting Fortune article)** | MPH | 1632 | 9 | 5 |
| 8/8/22 | Bummed that I missed Market Days over monkeypox fears, but glad I got my first vaccine dose this morning ✨  Still can't believe how much we're botching our response to this. Going to become a nightmare when schools reopen | PhD | 10100 | 21 | 0 |
| 8/8/22 | Start of the school year: 1) totally disregarded Covid-19, no measure whatsoever 2) monkeypox, but it’s MSM only so let it rip (until endemic) 3) hello swine flu 2022 🤯! So happy that COVID-19 “boosted” our immune systems!   https://cdc.gov/flu/swineflu/spotlights/first-human-infection-2022.htm… | PhD Professor Chemistry | 2703 | 6 | 1 |
| 8/8/22 | Cruz and GQP are neo-Nazis. They keep telling MAGAts that Dems are pedophiles to stoke violent Brownshirt attacks on LGBTQ ppl, teachers they smeared as "groomers." Lies like this + back-to-school monkeypox outbreak + CDC calling mpox "gay STD" = spark for pogroms, insurgent war. | PhD | 4673 | 3 | 1 |
| 8/8/22 | Just a mom preparing for another first day of school amidst an ongoing pandemic and a new one starting. It’s ok. We’re fine. #BacktoSchool2022 #WearAMask #covid #monkeypox | Doctor Researcher | 670 | 6 | 0 |
| 8/7/22 | The "monkeypox isn't serious. Nobody is dying so there's no need to shut down jobs or schools or events because of it" is happening and I...*deep negeo spiritual sigh* Just because something isn't deadly (most of the time) doesn't mean we should let it run wild. | Epidemiolgist | 1089 | 1 | 0 |
| 8/7/22 | My concern about monkeypox is colleges and schools this coming fall season. Dorm rooms are tricky with the traffic in and out. And If you live on campus you know it was parties and people having relations on campus. It could possibly be a major spread and the presidents | MS Adult Education | 792 | 2 | 1 |
| 8/6/22 | The U.S. is exhausting. The multiple ways this country tries to kill you in a day, especially if you’re Black, is exhausting. Now we talking #monkeypox like we have learned no lessons from 80s or the last two years. Tired. Starting the school year off with #Covid_19 & #monkeypox | PhD Professor Educational Policy | 39900 | 16 | 1 |
| 8/6/22 | Another example of why we need Monkeypox prevention guidance at nurseries & schools -> it’s mainly spread by close skin-to-skin contact. (And this is just basic prevention -> I don’t think anyone wants their child coming home with this disease) | PhD Professor Global Public Health | 319100 | 132 | 78 |
| 8/5/22 | So. What. Is. The. Plan?  I’m looking at all of you university and college and public school managers who are busy taping your back-to-school videos right now.  Shouldn’t you be figuring out what to do when monkeypox rips through your instituions ? | PhD Professor Engineering | 7153 | 75 | 21 |
| 8/5/22 | Female daycare worker has monkeypox. Many children exposed & MPX often more severe in kids. But go ahead & start schools with no significant protections because we've got to pretend everything's just fine . . . **(deleted tweet)** | PhD | 405 | 2 | 3 |
| 8/4/22 | So we really just collectively ignoring #Monkeypox? I really don't see many people talking about it or preparing for it.   Not paranoid about it but not ready for shorter days, hurricane season in Louisiana, and two pandemics with school starting in 3 weeks with no prep in sight. | PhD Student | 1208 | 2 | 0 |
| 8/4/22 | Sweet, can't wait to see what happens with this highly contagious virus when school starts back up.  As Monkeypox Spreads, U.S. Declares a Health Emergency | Professor Ecologist | 1628 | 1 | 0 |
| 8/4/22 | Off TOP the distribution of the monkeypox vaccine in Louisiana DURING PEAK FUCKING HURRICANE/EVACUATION SEASON AT THAT needs to be  1. Healthcare providers 2. First responders 3. The ELDERLY (esp. in nursing homes) 4. Teachers (school starts in 3 weeks & babies cant have it) | MPH | 17700 | 3 | 0 |
| 8/4/22 | Florida is a laboratory of Fascism. DeSantis is fueling it by targeting LGBTQ ppl and women. There will be soaring violence, esp w/ GQP using mpox as an accelerant, smearing teachers as 🏳️‍🌈 "groomers" just as school resumes, superspreading mpox among kids, as  @BarnabyBecky1  said. | PhD (anonymous) | 4673 | 11 | 6 |
| 8/4/22 | Let's recap the past three starts of the school year: August 2020- COVID. August 2021- COVID. August 2022- COVID+Monkeypox.  We are tired. | Ed.D | 6964 | 8 | 3 |
| 8/3/22 | How are schools, colleges, daycares and workplaces going to stay open with uncontrolled monkeypox and Covid spread? 🦠 🙈 🤷‍♀️ | Doctor (anonymous) | 10400 | 53 | 9 |
| 8/3/22 | There are now 96 confirmed cases of #monkeypox in children. 25 of those between the ages of 0-4. There is no vaccine available for children. School is starting with no protective strategies in place. What's the plan?  @POTUS @CDCgov @FLOTUS @VP https://worldhealthorg.shinyapps.io/mpx_global/#33_Case_profile_(overall) | MA Education | 6147 | 5 | 3 |
| 8/2/22 | Uhm so what IS the plan for monkeypox mitigation in schools? Just heard of first semi local probable case. Parents are concerned, they aren’t buying the “risk is low” this time around because we learned the hard way last time. Seriously, what. Is. The. Plan.  @teamMiPASS | PhD Microbiology | 1602 | 12 | 5 |
| 8/2/22 | More Monkeypox Cases Found In Kids As School Return Looms 🦠🙈 This isn’t going to be a fun school year. | Scientist | 14300 | 16 | 8 |
| 8/2/22 | Public health officials need to get ahead of this issue & imminent school return -> how do we have appropriate guidance & hygiene measures so we don’t see outbreaks among children in schools? Monkeypox spreads mainly through prolonged skin-to-skin contact | PhD Professor Global Public Health | 319100 | 142 | 84 |
| 8/2/22 | Hey @JCPSKY we're a week away from school starting, and parents have no information about how the district is addressing the threat of monkeypox. What's the plan, folks? It's been declared an emergency in two states already. | PhD | 15200 | 8 | 1 |
| 8/2/22 | Great thread on the need for a robust back-to-school plan to keep school communities safe in the time of #COVID19   Now, let’s add guidance on how to address #MonkeyPox because this is a growing, urgent concern in the U.S. right now. | PhD | 6715 | 5 | 0 |
| 8/2/22 | Monkeypox and covid are going to make this coming school year one of the toughest ever. The biggest issue is whether there will be reasonable PH measures in schools to protect children. | PhD (anonymous) | 597 | 36 | 11 |
| 8/2/22 | Once the school year starts monkey pox is going to explode rapidly! Kids are germ incubators and love touching everything and each other. | PhD Student | 1547 | 4 | 0 |
| 8/2/22 | New world record — More than 1,000 daily #monkeypox cases (7 days average). Those who said #MPXV would fizzle out soon are plain wrong. This fall school year will need radically new / more safety mitigations.   Figure by  @Antonio_Caramia | Epidemiolgist, ScD | 783100 | 3,366 | 1,818 |
| 8/1/22 | The spread of monkeypox is similar to what we see with viral meningitis. VM can spread by people living together: in school dormitories, households, shelters, and prisons. These clusters are self contained, so public health can come in, test, isolate, treat, and vaccinate. | HIV Researcher | 9375 | 11 | 2 |
| 8/1/22 | 8) Again, kids are much more vulnerable than adults for severe #monkeypox disease. While CDC says kids <8 are at high risk, the DHS report says kids <10 years old have been more frequently affected. Let’s take care to protect kids please! 🙏  https://dhs.gov/sites/default/files/2022-07/22_0712_st_monkeypox_mql.pdf **(comment)** | Epidemiolgist, ScD | 783100 | 1,042 | 425 |
| 8/1/22 | It is National Immunization Month, if you haven't had your 4th COVID shot, get it before school starts!and if you can, get a monkeypox vaccine. A grain of caution is much better than a pound of regret. | MPH Student | 998 | 8 | 1 |
| 7/31/22 | Society needs to be prepared for when today's minors sue parents, teachers, school districts, county public health offices, federal leaders, and others for knowingly exposing them to dangerous and deadly diseases. COVID is not over. Monkeypox is here. | PhD Professor History | 2748 | 1 | 0 |
| 7/31/22 | What the hell are we doing? Pretending all is well and monkeypox will just go away and not spread through schools and daycares?   If you’re in a position of power and you’re not advocating elimination before it’s too late, you have no ethical claim to authority. | PhD Student | 11400 | 51 | 18 |
| 7/30/22 | “Kids, constantly interacting at schools and daycare centres, may be especially vulnerable…if monkeypox were to start spreading in child-centric settings, it could be hard to contain.” 🦠🙈 | Scientist | 14300 | 9 | 1 |
| 7/30/22 | Talk to me about #UVC and #FarUV… what would a school need to cut the risk of MPX transmission in classrooms? Something that could sanitize the space between classes/during recess? Something that’s safe for use throughout the school day??  Give me hope, plz. #monkeypox  @PTF_org | MPH Student | 304 | 20 | 6 |
| 7/29/22 | Tried to schedule a vax appt for myself and my kids. Was told we couldn't be vaxxed for monkey pox because we aren't gay men... Only "at risk" populations are being vaxxed rn. Apparently immunocompromised and school children are not a priority. | Scientist | 614 | 5 | 0 |
| 7/28/22 | I wrote this in May,  @Surgeon_General    If I could see it, surely you could. Why did a ramp up not occur, and why are we still short? Once this hits schools we are toast.   #monkeypox | PhD | 2744 | 21 | 8 |
| 7/27/22 | So between monkey pox and the new COVID strain, we’re extra fucked for fall when schools, especially universities with large on-campus housing populations, return right? Go ahead and make that syllabus fully transferable to online teaching babes. | PhD Professor Gender Studies | 41600 | 41,600 | 6,477 |
| 7/27/22 | A lot of unknowns about the Monkeypox + Covid combination on global scale. Locally, one thing I am sure of: NOTHING is being done in schools in anticipation of the unknown and in response to the known. | PhD Humanities | 544 | 8 | 3 |
| 7/25/22 | I feel like we’re back in early 2020 with the way they’re treating monkeypox. Most people don’t even realize it can spread simply by touching something someone who has it touched or animals   And schools and colleges will be reopening in a few weeks 🥴🥴🥴😩😩 | PhD Agricultural Communication, Education/ Leadership | 1550 | 9 | 0 |
| 7/25/22 | We have to be prepared to counter this narrative when it erupts. The current “groomer” & anti-education rhetoric are going to collide with a potential monkeypox outbreak in schools to be a perfect storm of moral panic. | Doctor (anonymous) | 4040 | 6 | 0 |
| 7/24/22 | So...we just gonna open up the schools and colleges with no monkeypox OR covid safeties? Someone please let me off this train. | PhD Mathmatician | 5088 | 54 | 3 |
| 7/24/22 | What happens if someone gets COVID-19 and monkeypox at the same time? Do we know?   It would be irresponsible and exceptionally dangerous to start all in person school next month without knowing. | PhD Communications | 1642 | 9 | 0 |
| 7/24/22 | Resending this short reel on Monkey Pox I stole from Snark Tank. Most important point is that this is NOT an STD, and it could explode just in time for school! | PhD Chemist | 808 | 1 | 0 |
| 7/23/22 | Was just talking about this w/ a friend today: Covid, monkeypox, polio. School starts back for us at beginning of August. #Hellscape | Epidemiolgist | 1321 | 1 | 0 |
| 7/23/22 | When monkeypox gets into schools and daycares and gyms and workplaces and college campuses and y’all have to isolate and sanitize every piece of fabric in the building, just remember it was because no one cared enough about gay men. Sounds familiar… | Professor English | 6355 | 157 | 33 |
| 7/23/22 | When this spreads to the schools — and it will spread to the schools — will the crazies blame the LGBT community for Monkeypox? The  @whitehouse  and  @CDCgov  haven’t been forceful enough on combatting the hate that will@occur on this,  @POTUS . | Doctor | 1770 | 3 | 0 |
| 7/23/22 | Continuing to script monkeypox as a disease only affecting gay men is stigmatizing, preventing officials and the public from taking this seriously, and simply wrong.  When school resumes, this becomes bigger problem. Swift action has apparently eluded govs & Public Health. | PhD Student | 11400 | 42 | 14 |
| 7/23/22 | You’re frustrated that transportation and supply chains are a mess now. Just wait.   Politicians & public health have squandered the critical time to eliminate monkeypox. Sound familiar?  When school resumes it will take off. Kids can’t be vaxxed. Vax is in short supply. | PhD Student | 11400 | 20 | 5 |
| 7/23/22 | And the new school year didn't even start yet. Teachers going into battle every year with covid, staff shortages, mass shooters and now monkey pox.  JFC! | PhD Student (anonymous) | 8150 | 5 | 2 |
| 7/23/22 | “We won’t see #monkeypox in children” they said. Now have confirmed pediatric cases. They are linking everything to gay men, but community spread is here. Claiming monkeypox is like an STI was the worst messaging ever. School starts in a few weeks… | PhD Professor Economics | 34000 | 139 | 65 |
| 7/22/22 | Every school (K-12 and colleges) should be making plans for what they are going to do when/if monkeypox comes to their campus… | PhD Student (anonymous) | 24600 | 18 | 4 |
| 7/21/22 | When monkeypox spreads to the schools, they will absolutely create messaging of child molestation and grooming to target LGBTQ people when that monkeypox is not an STI/STD and the current messaging is absolute insanity. | MPH | 17700 | 75 | 30 |
| 7/21/22 | Oh no when monkeypox hits schools they're going to use it to force LGBTQ+ teachers out. Fuck | PhD Student | 48600 | 56 | 6 |
| 7/21/22 | I honestly cannot imagine that it wont spread in schools given that no one seems to be doing anything to stop the spread. I do not at all understand how people can say otherwise. Unless they think men who have sex with men are never parents? Or they do not understand transmission **(in a thread discussing Mpox) (comment)** | ScD Professor | 125000 | 90 | 14 |
| 7/20/22 | When monkeypox shows up in your kid’s schools or daycare this fall will you: a) be notified? b) keep sending your kid? c) keep your kid home? d) feel reassured when officials say “it’s mild”? e) feel reassured when told that “infection builds immunity”? | PhD Student | 11400 | 110 | 33 |
| 7/20/22 | Between COVID and Monkey Pox, schools (K-12 and colleges) should be making plans now for what they are going to do when/if they have serious outbreaks when schools reopen in a bit more than a month.  Better to have a plan and not wind up needing it, than to have no plan. | PhD Student (anonymous) | 24600 | 40 | 12 |
| 7/20/22 | Trying to dodge Covid & monkeypox should make for an interesting school year 🥴 | Doctor | 12800 | 52 | 12 |
| 7/20/22 | Quebec parents of elementary school children will likely find 3 boxes of tissues on their school supply lists this year, rather than 2, and this will be the extent of Covid preparedness offered by institutions (paid for by parents). And monkeypox? What's that? WAKE UP. | PhD Humanities | 544 | 4 | 0 |
| 7/20/22 | Once monkeypox gets into the schools and pre-ks and daycares?   Boyyyyyyyyy. | MPH | 17700 | 6 | 2 |
| 7/18/22 | Is there any reason to think that monkeypox won't be spreading in schools, like, soon-ish? | Epidemiologist | 4367 | 654 | 104 |
| 7/16/22 | Monkeypox is spread through close skin-to-sking contact. Perhaps University and High School wrestling programs need to be suspended temporarily… | Parasitologist | 6409 | 2 | 0 |
| 7/13/22 | The longer monkeypox circulates, the higher the likelihood of a school contact like this initiating an outbreak in another highly connected group.  London school sends reception class home over Monkeypox fears https://standard.co.uk/news/london/monkeypox-grand-avenue-school-surbiton-vaccine-ukhsa-b1012005.html | Paleovirology | 36200 | 271 | 111 |
| 7/2/22 | Monkeypox is largely spread by skin-to-skin contact (there are also other methods). Perhaps high school/university wrestling programs should be temporarily suspended… | Parasitologist | 6409 | 2 | 1 |
| 6/28/22 | Let's dispel the myth that monkeypox is mostly an STD. Here is a case in a primary school aged child in the Netherlands. | Epidemiologist | 15200 | 28 | 9 |
| 6/8/22 | Isn't monkeypox mostly fatal in children? If so, then "no deaths so far despite 1000 (adult) cases" =/= "it's mild". We need to prevent it from getting into schools. | PhD Professor Evolutionary Biology | 80200 | 415 | 65 |
| 5/21/22 | Don’t ever send your child to public school 🏫 during a SARS and Monkey Pox dual pandemic. I know this his hard to understand. But permanently disabling and disfiguring your kid is not a good idea 🦠 | Doctor (anonymous) | 10400 | 1 | 0 |
| 5/20/22 | NEXT: 15 days to slow the spread. #monkeypox   Cruises and flights canceled. Colleges and universities sending students home. Public schools closing.  Offices asking people to telecommute.  Concerts, parades, festivals and sporting events postponed. 🙄 | PhD | 565 | 1 | 0 |
| 5/20/22 | With monkey pox SARS and hepatitis. Maybe ending school a month early would be in the best interest of the kids and the whole fucking world | Doctor (anonymous) | 10400 | 5 | 0 |
| 5/18/22 | Hey  @VCHhealthcare  &  @CDCofBC , here's an off-ramp: how about implementing airborne protections *BECAUSE OF MONKEYPOX*. We don't want it in hospitals and schools, hence N95s, ventilation, filtration. We don't even need to talk about that other virus which is totally not airborne. | Professor Economics | 3076 | 7 | 3 |
| **JD** |  |  |  |  |  |
| **Date** | **Tweet** | **Author Credentials** | **Follower Count** | **Likes** | **Retweets** |
| 8/30/22 | Prediction: We’re going to have #monkeypox spreading in schools because the CDC couldn’t be bothered to mount an effective response to the initial outbreak.  Enjoy. | JD | 2394 | 6 | 0 |
| 8/22/22 | Monkeypox is Diet Smallpox  It’s very serious It’s very contagious  It’s not a STD  It can easily run through a school. | JD | 3164 | 5 | 0 |
| 8/22/22 | Is anyone doing anything about monkeypox and back to school? Or we cool just like that? Because I'm pretty sure hugging is going to be on most people's agenda on campuses everywhere. | JD Professor | 44500 | 368 | 52 |
| 8/11/22 | Monkey pox is on track to become a completely uncontrollable disaster because of all this nonsense. And it’s only going to get worse now that schools are starting up again. It’s infuriating. | JD Student | 4071 | 14 | 0 |
| 8/4/22 | And the CDC wants to just get rid of COVID protective measures right before school starts (which would also limit the transmission of monkeypox) | JD | 990 | 2 | 0 |
| 8/4/22 | COVID is ever present, monkeypox is on the rise as well as polio, yea that’s right polio….   We are also entering the worst of flu season AND SCHOOL IS ABOUT TO START!   Wash your hands. wear your mask, keep sanitizer, social distance as best as you can.  😭 | JD Professor | 4423 | 3 | 0 |
| 8/2/22 | Listen to people who have been proven correct, like Dr. Eric Feigl-Ding, not the minimizers who mistake wishful thinking for science. Monkey Pox is not a joke and will not fizzle out. And schools are about to start. 🤦‍♀️ | JD Retired | 515 | 3 | 0 |
| 7/24/22 | School is about to start & the research has said in the areas of the world that have had monkeypox outbreaks, children will get it worst than adults. We’ve already had adults share how painfully bad their monkeypox experiences have been. We’ve just seen the exact thing w/ covid. | JD | 8160 | 3 | 5 |
| 7/24/22 | Unfortunately not seeing the pics of monkeypox isn’t going to make it go away as the cases multiply. The country is at 2500 cases as of yesterday. And there’s no contingency plans with school about to start. This ain’t chickenpox. You need to see it to know what to look for | JD | 8160 | 12 | 7 |
